# The impact of loneliness on healthcare costs and service utilisation and the cost-effectiveness of loneliness interventions: a systematic review

**DOI:** 10.1101/2024.12.14.24318998

**Authors:** Sharon Eager, Helen Baldwin, Paul McCrone, David McDaid, Theodora Stefanidou, Prisha Shah, Stephen Jeffreys, Antonio Rojas-Garcia, Ruby Jarvis, Beverley Chipp, Brynmor Lloyd-Evans, Alexandra Pitman, Phoebe Barnett, Maria Ana Matias, Sonia Johnson

## Abstract

**Introduction:** Loneliness is an unpleasant, subjective experience that can affect people at all stages of life. Given its association with a range of physical and mental health problems, it is important to assess the costs of loneliness to the healthcare system. The current study aimed to: (i) review literature on the health and social care impacts of loneliness, and (ii) review economic evaluations of interventions to address loneliness.

**Methods:** We conducted a systematic review of studies published from 2008 to 2023 by searching five bibliographic databases, two sources of grey literature, and reference lists of relevant systematic reviews. Studies estimating health and social care cost and expenditure, and studies on health resource utilisation were included to assess the impact of loneliness on the health system. Return on investment, social return on investment, and cost-effectiveness evaluations were included to assess the economic impact of loneliness interventions. Studies that reported either mental or physical health outcomes were included. No limitations were placed on inclusion for populations, age groups, or language. We conducted quality appraisal and narrative synthesis of results.

**Results:** We included 39 eligible studies: Five estimated the healthcare cost or expenditure of loneliness, 22 reported healthcare resource use, and 18 were economic evaluations of interventions. Most studies focused on older adult populations. Findings relating to the cost/expenditure of loneliness and service use were inconsistent and varied: some studies reported excess healthcare costs/expenditure and service use associated with loneliness, while others found lower costs/expenditure and service use associated with loneliness. Economic evaluation studies covered a wide range of different intervention types. They generally indicated that loneliness interventions can be cost-effective, but were less likely to be cost-saving and not consistently effective in reducing loneliness.

**Discussion:** Inconsistent and varied findings on the impact of loneliness on the healthcare system and on economic evaluations of loneliness interventions meant it was difficult to synthesise the results. Therefore, we cannot derive confident conclusions about the impact of loneliness on costs/expenditure related to the health and social care system from this review. Future research should prioritise studies relating to social care, the direct healthcare costs of loneliness, and randomised controlled trials with long-term follow-ups to appropriately address these evidence gaps. Research on younger populations should also be considered a priority.

## Introduction

Loneliness is an unpleasant, personal experience that arises when someone feels their social relationships are deficient in terms of quality and/or quantity [1,2]. Loneliness is closely related to social isolation, which refers to someone’s objective number of social contacts and interactions [3]. Although these concepts are related, the subjective nature of loneliness distinguishes it from objective social isolation [4]. While loneliness has been thought of previously as an issue primarily affecting older adults, recent studies have demonstrated a U-shaped distribution of loneliness, with its prevalence particularly pronounced in late adolescence/early adulthood [5–7].

Loneliness is associated with a variety of physical and mental health problems [8,9]. Among mental health outcomes, there is most consistent evidence to support an association with depression [10,11]. Formal synthesis of the evidence demonstrates that lonely people have around double the odds of developing depression compared to non-lonely people [12]. Loneliness is also a predictor of the subsequent development of anxiety disorders [13], and is associated with more severe depression and anxiety symptoms among people with existing mental health problems [14]. It also predicts suicidal ideation, suicide attempts and suicide [15,16]. In terms of physical health, loneliness has been linked to cardiovascular problems, including higher blood pressure, lower cardiac output, and cardiovascular disease compared to non-lonely participants [17–19]. Chronic loneliness has been linked with higher stroke risk [20]. An association has been demonstrated between loneliness and risk of dementia, characterised by more rapid cognitive decline in older age [21,22]. Loneliness is also associated with an elevated risk of premature mortality [23,24].

Over the last decade, there has been increased policy interest in addressing loneliness. For instance, in 2018 the UK Prime Minister launched a cross-sectoral strategy to tackle the issue in England [25]. The negative health outcomes associated with loneliness and the increase in loneliness and isolation reported during the COVID-19 pandemic [26] have accelerated a rapid increase in research and public interest in loneliness and its societal impact [27]. Thus a case has been made that loneliness should be considered a pressing public health issue [23,28]. An important aspect of understanding the consequences of loneliness is to assess its economic impact. Due to the adverse associations between loneliness and mental and physical health outcomes, an association might be anticipated between loneliness and variations in patterns of health and social care service use [29].

There have been recent efforts to summarise evidence on the healthcare use and costs associated with loneliness, but these evidence summaries have had their limitations. They include a systematic review of the economic costs of loneliness and the cost-effectiveness of loneliness interventions [29] and a systematic review regarding the association between loneliness and health and social care use in older adults[30]. The former review, by Mihalopolous and colleagues [29], identified 12 studies of samples across all age ranges, including cost-of-illness studies, economic evaluations, return on investment (ROI) studies, and social return on investment (SROI) studies. The authors found some indications that loneliness may be associated with additional costs and that loneliness interventions may be cost-effective, but they noted inconsistency in the included studies and concluded that comparability across studies was limited. The latter review, by Smith and Victor [30], identified 32 heterogeneous studies (solely focused on older adult populations), and concluded that their findings were insufficient to determine whether loneliness was associated with changes in health and social care use among older people. Both reviews highlighted a paucity of evidence on this topic, especially in relation to general adult and younger populations.

An important limitation of these reviews was that they either only examined changes in the utilisation of services, or summarised evidence on health and social care costs, but did not consider these together. Another limitation was that these reviews did not look at the relative impacts on different population groups across the life course, such as young people, working age populations, and older adults. They also did not consider evidence on loneliness published since the onset of the COVID pandemic. Given these limitations and the need for an updated and comprehensive summary of this literature, we undertook a systematic review of the health and social care costs and expenditure of loneliness, the association between loneliness and health and social care use, and the economic value of interventions aiming to ameliorate loneliness in both the general population and specific at-risk groups. Our aims were:

1. To review available literature on the healthcare impact of loneliness, including studies estimating the health and social care cost and expenditure of loneliness, as well as studies looking at the association between loneliness and health and social care resource use;
2. To review available economic evaluations of interventions whose primary purpose was to address loneliness.

## Methods

### Study design

We designed our review strategy as a team composed of academics, researchers with lived experience, and clinical academics. We pre-registered the protocol for this systematic review (PROSPERO Protocol Registration Number: CRD42023402725) and adhered to the principles of the Preferred Reporting Items for Systematic Reviews (PRISMA) guidelines [31].

### Search strategy

We conducted systematic database searches in MEDLINE, EMBASE, PsycINFO (using Ovid), CINAHL and EconLit (using EBSCOhost) using relevant keyword and subject heading searches (see Supplementary file A) for studies published between January 2008 and March 2023. We conducted grey literature searches via Google Scholar search and EThOS. We also hand searched the reference lists of any relevant systematic reviews identified for potentially relevant studies.

### Screening

We de-duplicated all database search results using Endnote version x9 [32] and imported these into Rayyan [33] for screening. We conducted title and abstract screening independently, involving seven researchers in this task (SE, TS, PM, DM, PS, SJe, and ARG). Each record was independently double-screened by two researchers for consistency. All full texts were screened in the same way. Any discrepancies between researchers’ decisions were resolved through discussion with the wider research team.

Inclusion and exclusion criteria We included the following studies:

1. Studies primarily reporting the impact of loneliness on health and social care. These included studies estimating the health and social care cost and/or expenditure associated with loneliness, and studies on health and social care service utilisation (including primary care use, visits to secondary care specialists, and inpatient care, with and without costs reported).
2. Economic evaluations of intervention studies on the prevention or alleviation of loneliness, including ROI, SROI, and cost-effectiveness evaluations.

We included peer-reviewed scientific papers, published reports, and theses that related to either mental and/or physical health outcomes. There were no limitations placed upon included populations, age groups, or language of publication. We included studies published since 2008 to reflect the increased prevalence of loneliness research published in the last 15 years [27]. We excluded non-systematic narrative reviews, qualitative studies, and book chapters.

### Data extraction

Eight researchers conducted data extraction and quality appraisal (SE, HB, TS, RJ, PS, DM, PM, ARG). All extractions and quality appraisals were cross-checked for accuracy by one researcher from a sub-set of the team (SE, HB, DM, PM). Missing data were requested from study authors.

We extracted relevant data into a proforma including fields pre-agreed by the team. Data extraction for all studies included: bibliographic details, country, study aims, sample size, sample health status, sample demographics, study design, loneliness measures used, lived experience involvement, cost perspective, costing/pricing year and currency, time horizon of study, and author summary/conclusions. For studies estimating the cost/expenditure of loneliness and reporting health resource use, we additionally extracted direct and indirect costs/resource use outcomes and direct and indirect costs/resource use results. For economic evaluation studies, we additionally extracted information on the intervention and comparator, type of economic evaluation, main economic outcome measure, discount rate, types of costs measured, main outcomes, costs reported, and economic evaluation results.

### Quality appraisal

We used the Schnitzler et al. [34] quality appraisal checklist to assess the quality of evidence for studies estimating the cost/expenditure of loneliness and healthcare use. We used the Evers et al. [35] checklist to appraise the quality of these economic evaluation studies. For quality appraisal, each item within the corresponding checklist was weighted equally and we reported scores as a percentage of the total number of applicable items for each study. We had initially planned to use different checklists to assess quality, as reported in our protocol: namely, the Larg & Moss checklist [36], Drummond checklist [37], and Krlev checklist [38]. However, during the course of our study, Schnitzler and colleagues [34] published a checklist for the appraisal of cost-of-illness studies, which we found to be more appropriate for the studies retrieved in our search. We also identified that the Evers [35] checklist for economic evaluation studies was more consistent with the newly published Schnitzler checklist [34] and therefore elected to use this for the appraisal of economic evaluation and ROI studies, rather than the checklists described in our protocol.

### Evidence synthesis

We carried out a narrative synthesis of eligible studies [39]. This included developing a preliminary synthesis of findings from included studies, exploring potential relationships in the data, including comparisons of study quality, design, costing approach, population group and setting, as well as finally assessing the overall robustness of the synthesis.

To facilitate comparisons, all monetary values (excluding ROI and SROI values) have been adjusted for purchasing power parity (PPP) using the CCEMG–EPPI Centre Cost Converter version 1.7 [40]. The reference year for PPP adjustments was 2023 and our standard currency was USD. All PPP values are reported in brackets after originally reported figures.

## Results

### Study selection

Our searches identified 1,842 records, which were reduced to 1,194 studies after de-duplication. On screening these titles and abstracts, we identified 203 for full text review. From these, 39 studies were eligible for inclusion. These comprised five studies estimating the healthcare cost or expenditure of loneliness, 22 healthcare resource use studies, and 18 economic evaluations. Full details of the search and screening process are provided in the PRISMA flowchart in Figure 1. See Supplementary file B for a list of all papers excluded at full text screening, including exclusion reasons.

**Figure 1.**
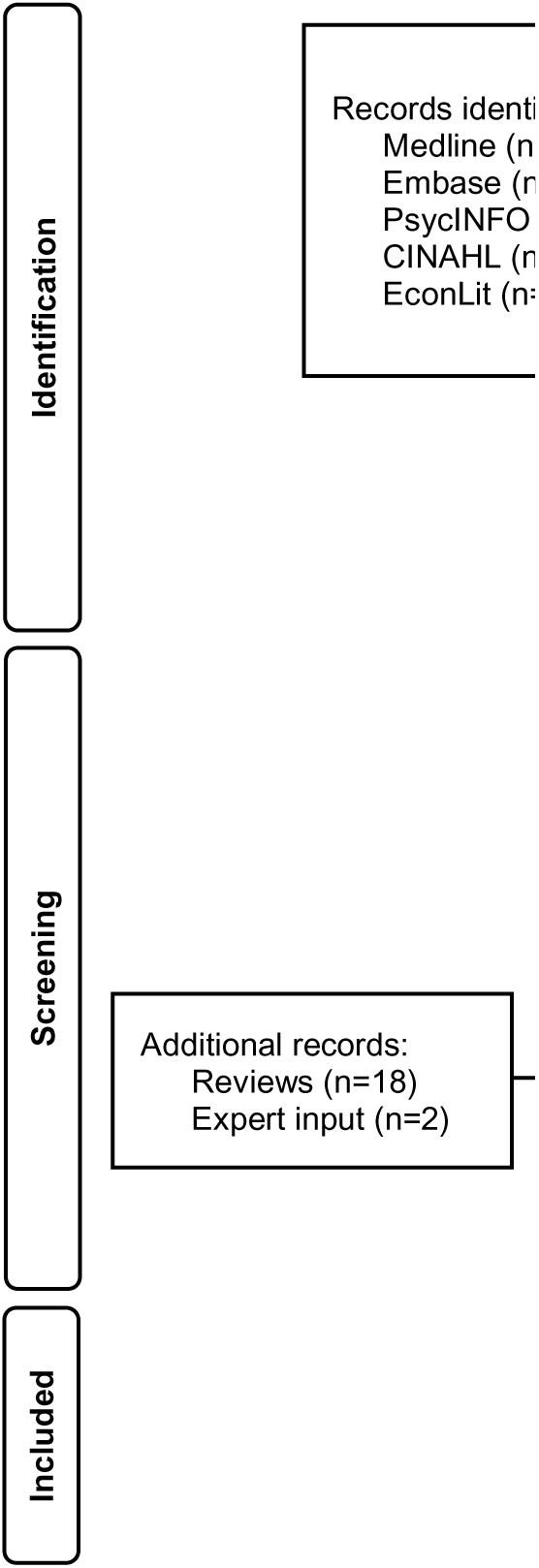
PRISMA flow diagram summarising the search.

### Quality of included studies

The quality of included studies was judged to be generally moderate to high. Of the 27 studies estimating the healthcare cost of loneliness and studies examining the association between loneliness and healthcare use that were assessed for quality using the Schnitzler checklist [34], 18 met between 60% and 77% of quality criteria. The remaining nine studies met between 52.6% and 41.7% of quality criteria. The criteria that were most often missed across studies related to whether sensitivity analyses were conducted, whether the time horizon of the study was justified, and whether ethical issues were discussed. Of the 18 economic evaluations that were assessed for quality using the Evers checklist [35], nine of these met between 81.3% and 94.4% of quality criteria. Seven studies met between 68.4% and 78.9% of quality criteria, with the remaining two studies meeting 58.8% and 44.4% of criteria respectively. The quality criteria these studies were rated lowest on most consistently related to whether they discussed ethical and distributional issues and whether they discussed the generalisability of findings. See Supplementary file C for full quality appraisal ratings for each study.

### Studies estimating the healthcare cost and expenditure of loneliness

Five studies examined the healthcare costs or expenditure associated with loneliness, of which two also examined aspects of social care (Table 1). Of the five studies in this category, three employed a cross-sectional design, one a longitudinal design, and one was a modelling study. Only two studies looked at the healthcare cost of loneliness while the remaining studies examined expenditure. Four studies investigated the cost or expenditure of loneliness in older adults (65+), and the other examined healthcare expenditure in the general adult population. Three studies were from the USA, and one each was from the UK and the Netherlands. Three measured loneliness using the 3-item UCLA Loneliness Scale and one used the 11-item De Jong Gierveld Scale. None reported any public or patient involvement (PPI) or lived experience input in design, delivery, or interpretation.

**Table 1.** Summary of studies estimating the health and social care cost and expenditure of loneliness (n=5).

| *Lead author, publication year* | *Country; currency; costing year* | *Study aim(s)* | *Study design* | *Sample description; sample size* | *Loneliness measure; variables adjusted for in analysis* | *Results* | *Quality score using Schnitzler checklist [34]* |
| --- | --- | --- | --- | --- | --- | --- | --- |
| Barnes et al., 2022 [45]. | USA, USD; 2018-2019. | Examine the cumulative effect of loneliness and social isolation on various health outcomes (including emergency department visits, inpatient admissions, and total medical costs). | Cross-sectional survey. | Older adults (65+); mean age=76.5 years; 45% male, 55% female; N=6,994. | 3-item revised UCLA Loneliness Scale; socio-demographic and healthcare factors. | Overall mean medical cost for non-lonely participants was \$9,292 (PPP=\$11,107.89). Mean medical cost for participants who were lonely (but not socially isolated) was \$10,350 (PPP=\$12,372.65). Mean medical cost for both socially isolated and lonely participants was \$13,008 (PPP=\$15,550.09). The differences in medical cost for lonely participants (those who were lonely only and those who were both lonely and socially isolated) compared to non-lonely participants were not statistically significant. | 10/24 (41.7%) |
| Fulton & Jupp, 2015 [42]. | UK; GBP; 2010-2012. | Model the impact of loneliness on GP visits, A&E visits, unplanned admissions, residential care, depression, dementia, and physical inactivity, and provide an assessment of the potential cost to the public sector. | Modelling study drawing on published literature. | Older adults who report feeling lonely compared to average older population; not reported. | N/A; N/A. | Average cost associated with being chronically lonely to the public sector, based on increase in service usage, is £12,000 (PPP=\$23,032.63) over the medium term (15 years) per person, compared to those who are never lonely. Estimate based on additional cost of £150 (PPP=\$286.53) for GP visits, £27 (PPP=\$51.57) for A&E visits, and £56 (PPP=\$106.97) for unplanned admissions, all per person per annum. Additional costs considered in the model were increased likelihood of residential care, depression, dementia, and physical inactivity among lonely individuals. | 11/22 (50%) |
| Meisters et al., 2021 [43]. | Netherlands; EUR; 2017. | Investigate the relationship between loneliness and healthcare expenditure (total, GP, specialised, pharmaceutical, and mental healthcare spending) using a large, nationally representative, sample. | Three combined cross-sectional datasets. | General population of adults; Mean age=59.3 (SD= 16.9), 47.3% male, 52.7% female; N=341,376. | 11-item De Jong Gierveld scale; socio-demographic factors, self-perceived health, and psychological distress. | Loneliness was associated with increased mental healthcare expenditure (IRR for very severely lonely=1.31, CI=1.08-1.58, $p<0.05$), which translated to €340.2 million (PPP=\$525.55 million) mental healthcare expenditure (CI=314.7 to 365.8), or 10.3% of annual mental healthcare spending. Small increase in GP expenditure associated with very severe loneliness (IRR=1.08, 1.04-1.13, $p<0.05$). This corresponded to €5.8 | 14/22 (63.6%) |
| | | | | | | million (PPP=\$8.96 million) GP expenditure (CI=4.5 to 7.1), or 0.8% of total annual GP spending. Negative association between very severe loneliness and specialised care expenditure (IRR=0.88, CI=0.8 to 0.97, $p<0.05$), which corresponded to €449.8 million (PPP=\$694.86 million) lower specialised care expenditure (CI=-474.3 to -425.4), or 2% total annual spending. Small negative association between somewhat lonely individuals and total expenditure (IRR=0.96, CI=0.93 to 0.99, $p<0.05$). This translated to €435.4 million (PPP=\$672.62 million) lower total healthcare spending (CI=-494.8 to -376.1), or 1% of total population annual healthcare spending. | |
| Musich et al., 2022 [44]. | USA; USD; 2018 and 2019. | Investigate the association of physical activity levels with loneliness, social isolation, protective factors: resilience, purpose-in-life, and perception of aging; and the impact of these factors on | Cross-sectional survey. | Older adults (65+); 45.5% male, 54.6% female; N=6,652. | 3-item revised UCLA Loneliness Scale; socio-demographic factors and health status. | For participants who had moderate physical activity levels, those who were severely lonely had significantly higher annual healthcare expenditure (\$12,338; PPP=\$14,489.28) than those with low/no loneliness (\$9,154; PPP=\$10,750.11; $p=0.002$). For participants who had high and low physical activity levels, there were no significant differences in healthcare | 12/23 (52.2%) |

The study with the highest quality score (77%) was conducted in the USA [41]. This was a longitudinal assessment of annual Medicare (the main federal insurance scheme for older Americans) health and social care spending for 5,270 individuals aged 65 years and older. Mean monthly health spending was $1,024 (PPP=$1,350.68). Spending covered inpatient acute care, hospice care, post-hospitalisation nursing home care, outpatient services, and medical providers’ fees, including primary care, specialist care, occupational/physical therapies, and some home health services. Compared to non-lonely participants, those who were lonely represented significantly lower expenditure, $64 (PPP=$84.42) per month (*p*<0.001), or $768 (PPP=$1,013.01) per year. Overall mean monthly inpatient spending was $396 (PPP=$522.33), with lonely participants having $54 (PPP=$71.23) lower monthly inpatient spending (*p*<0.05) than non-lonely participants. Loneliness did not explain differences in expenditure for outpatient care or skilled nursing facility care (i.e., post-hospitalisation nursing home care).

In a study judged to be of moderate quality, data from prior studies sourced from within and external to the UK examining the cost of loneliness were used to model the cost of loneliness over 15 years among older adults in the UK [42]. In contrast to the findings of the above USA study [41], the UK authors [42] estimated the additional costs for an older person experiencing chronic loneliness as £12,000 (PPP=$23,032.63) higher than a non-lonely person over a 15-year period. Their model included estimates of the annual cost per person of increased primary care visits (£150; PPP=$286.53), emergency room visits (£27; PPP=$51.57), and unplanned admissions (£56; PPP=$106.97) for those reporting loneliness compared to non-lonely individuals. They also accounted for costs associated with an increased likelihood of lonely individuals entering local authority-funded residential care and experiencing health conditions (including depression, dementia, and physical inactivity). They estimated that 40% of these costs occurred within 5 years of an older person experiencing chronic loneliness.

The three remaining studies in this category presented cross-sectional analyses. In a Dutch study judged to be of moderate quality, multiple databases were combined to achieve a large (N=341,376), nationally representative sample of adults in the Netherlands[43]. The authors calculated expenditure figures based on health insurance spending in their sample, and then extrapolated this figure to the entire Dutch population. They found that when compared to non-lonely individuals, those who were rated as *somewhat lonely* represented a slightly lower total expenditure (IRR=0.96, CI=0.93-0.99, *p*<0.05), which translated to €435.4 million (CI=-€494.8 to -€376.1; PPP=$672.62 million) reduced total population spending, or 1% of total health expenditure. They also found lower specialised care expenditure for people who self-rated as *somewhat lonely* (IRR=0.94, CI=0.90-0.98, *p*<0.05) or as *very severely* lonely (IRR=0.88, CI=0.80-0.97, p<0.05). This corresponded to €449.8 million (CI=-€474.3 to - €425.2 million; PPP=$694.86 million) lower spending on specialised care, corresponding to 2% of total population specialised care spending. In contrast, there was an association between higher expenditure on GP visits and self-rating as *somewhat* (IRR=1.02, CI=1.01-1.04, *p*<0.05), *severely* (IRR=1.07, CI=1.04-1.10, p<0.05), and *very severely* (IRR=1.08, CI=1.04-1.13, *p*<0.05) *lonely*. This translated to €5.8 million (CI=€4.5-€7.1; PPP=$8.96 million) additional spending on GP visits, or 0.8% of total annual primary care expenditure. The authors also found an association between increased mental healthcare expenditure and self-rating as *somewhat lonely* (IRR=1.17, CI=1.04-1.33, *p*<0.05) or as *very severely lonely* (IRR=1.31, CI=1.08-1.58, *p*<0.05), corresponding to €340.2 million (CI=€314.7-€365.8; PPP=$525.55 million) extra mental healthcare spending, or 10.3% of annual mental healthcare expenditure. The authors did not observe a significant association between loneliness and pharmaceutical expenditure.

In another cross-sectional study from the USA, rated as moderate quality, the authors investigated how loneliness impacted on healthcare expenditure patterns according to level of physical activity in a sample of 6,652 older adults [44]. Participants who had moderate physical activity levels and who were rated as severely lonely had a mean annual healthcare expenditure of $12,338 (PPP=$14,489.28), while those who were rated as having low/no loneliness had significantly lower mean annual expenditure of $9,154 (PPP=$10,750.11; *p*=0.002). For participants with either high or low physical activity levels there were no significant differences in their healthcare expenditure according to whether they reported loneliness or not. Finally, in another USA cross-sectional study involving almost 7,000 older adults rated as low-moderate quality, no significant association was found between loneliness and increased total medical costs [45].

In summary, two studies looking at expenditure reported a significant association between loneliness and lower healthcare expenditure while one study reported higher expenditure for individuals with severe loneliness compared to no/low loneliness. In terms of costs, one study estimated excess healthcare cost associated with loneliness, and the remaining study did not report significant association between loneliness and healthcare costs.

### Healthcare resource use studies

The 22 studies evaluating the association between loneliness and health and social care use are summarised in Table 2. Where authors presented estimates as unadjusted, partially adjusted and/or fully adjusted associations, taking into account specified confounders, we report estimates from the final model only.

**Table 2.**
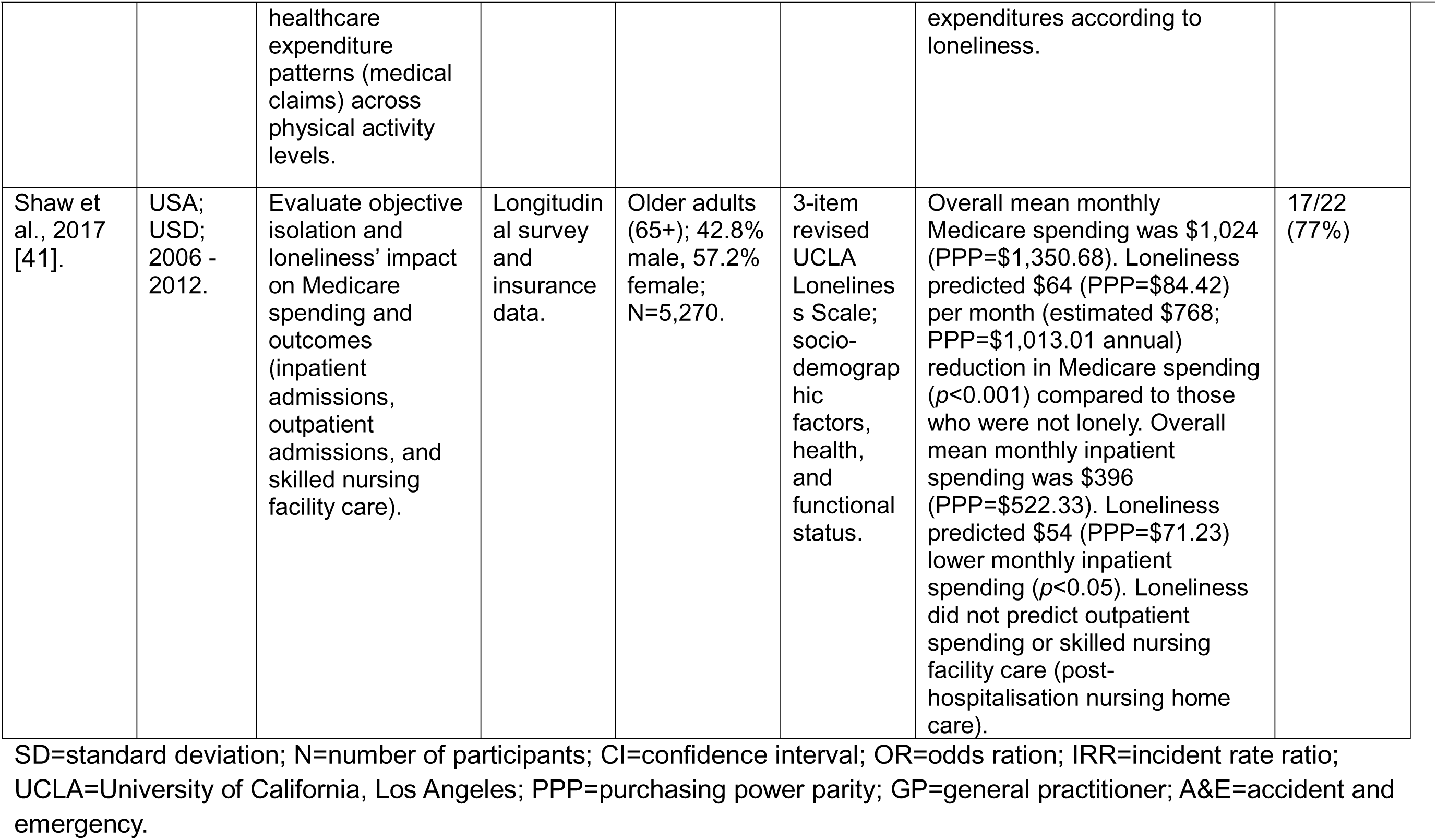
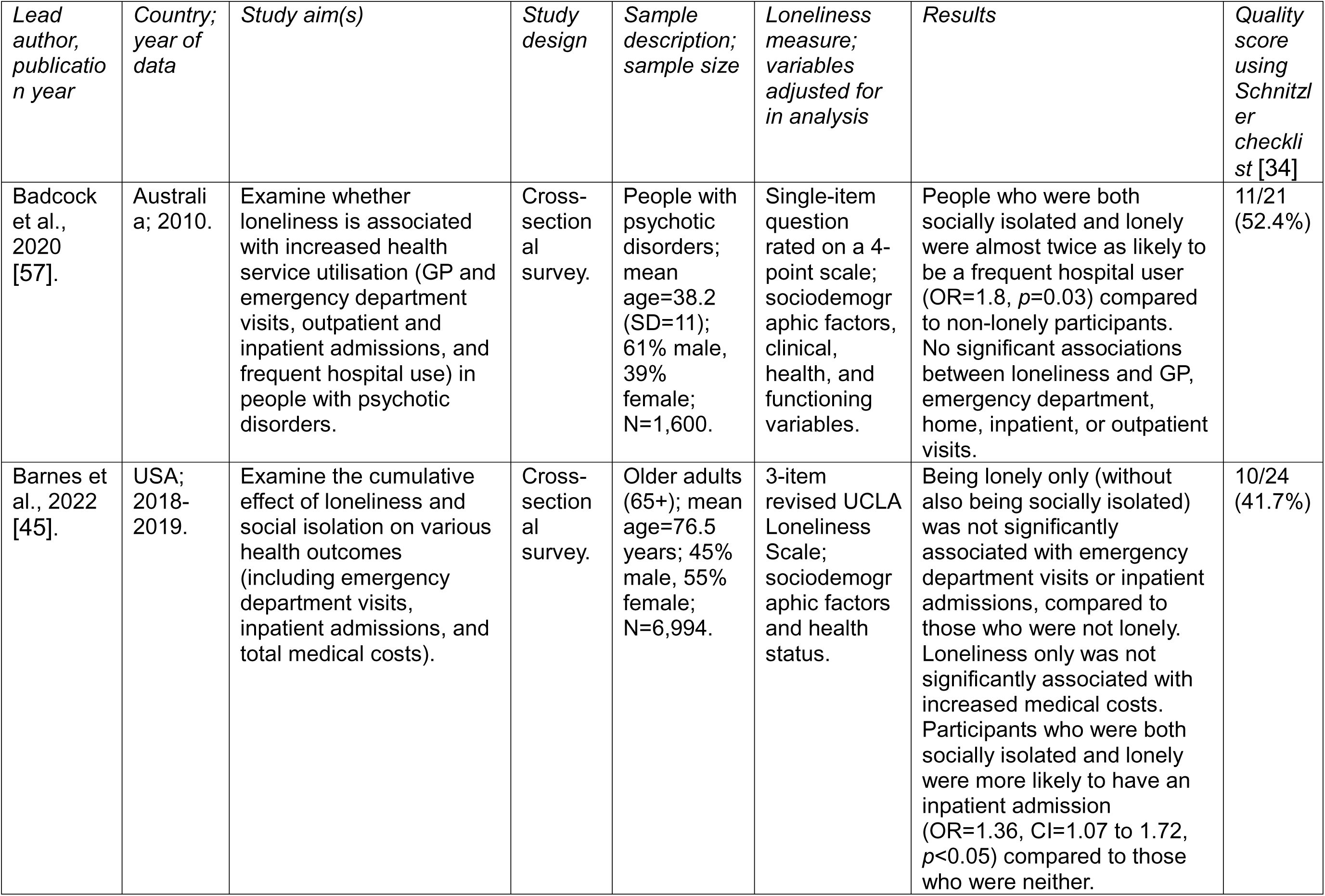
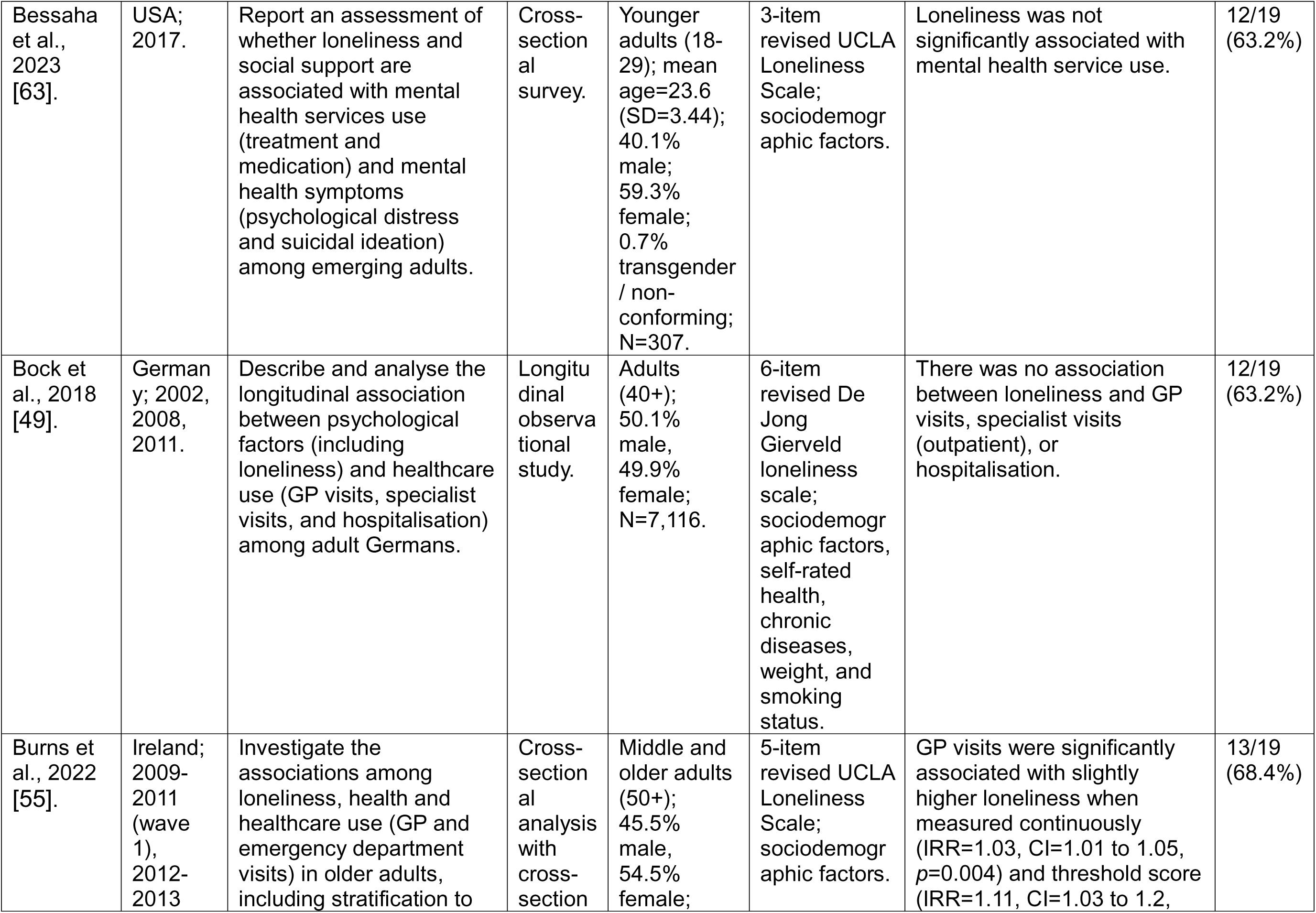
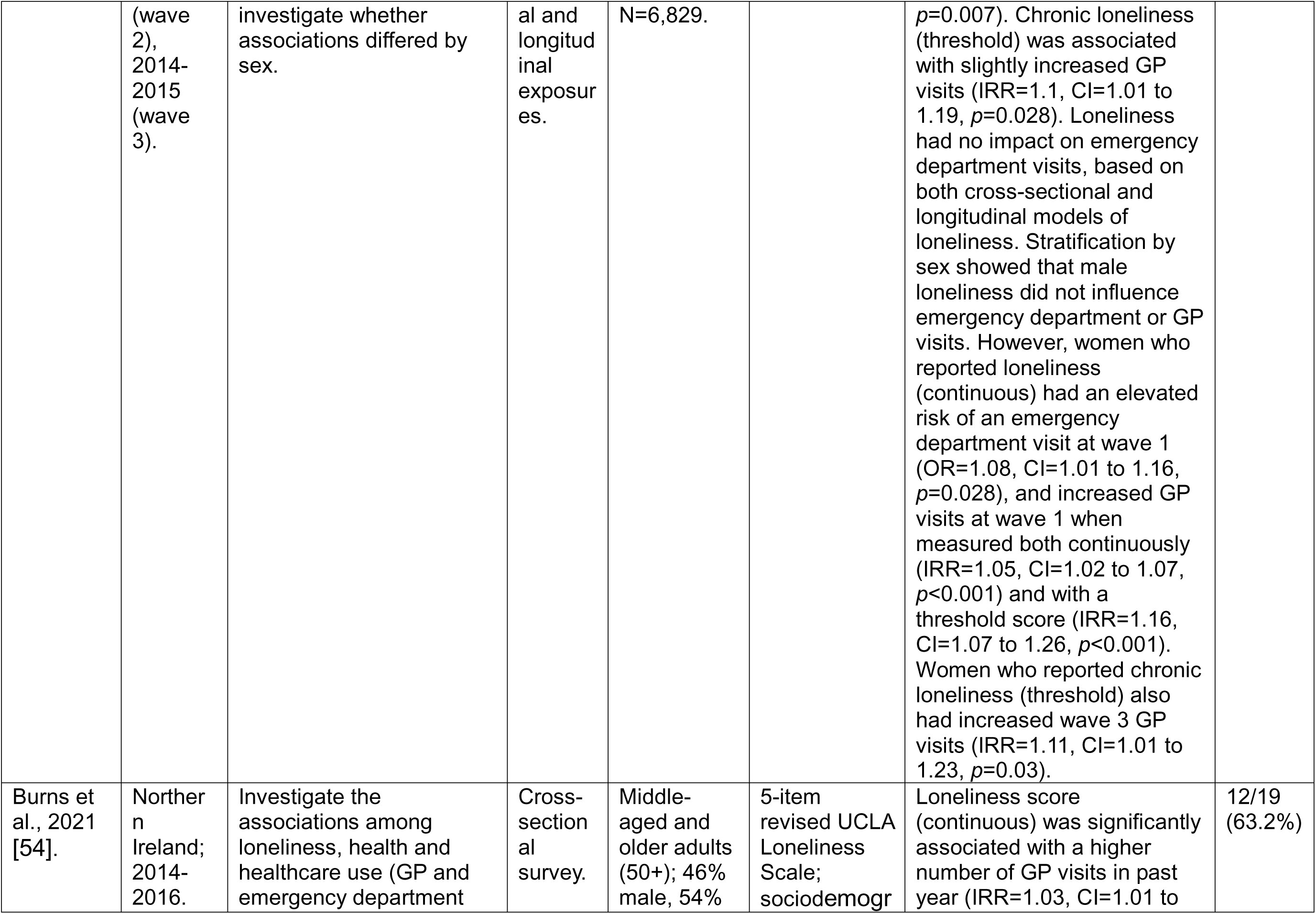
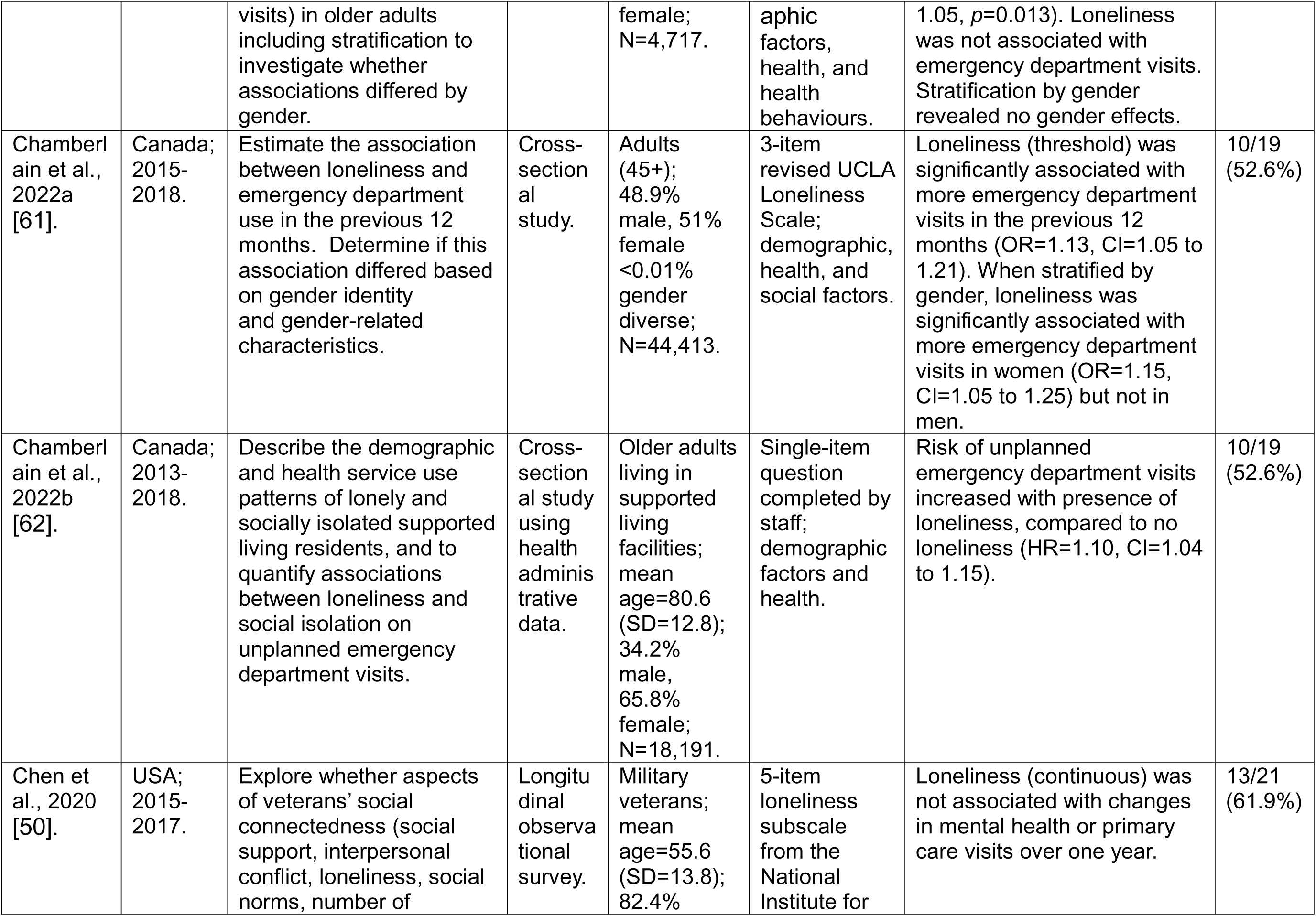
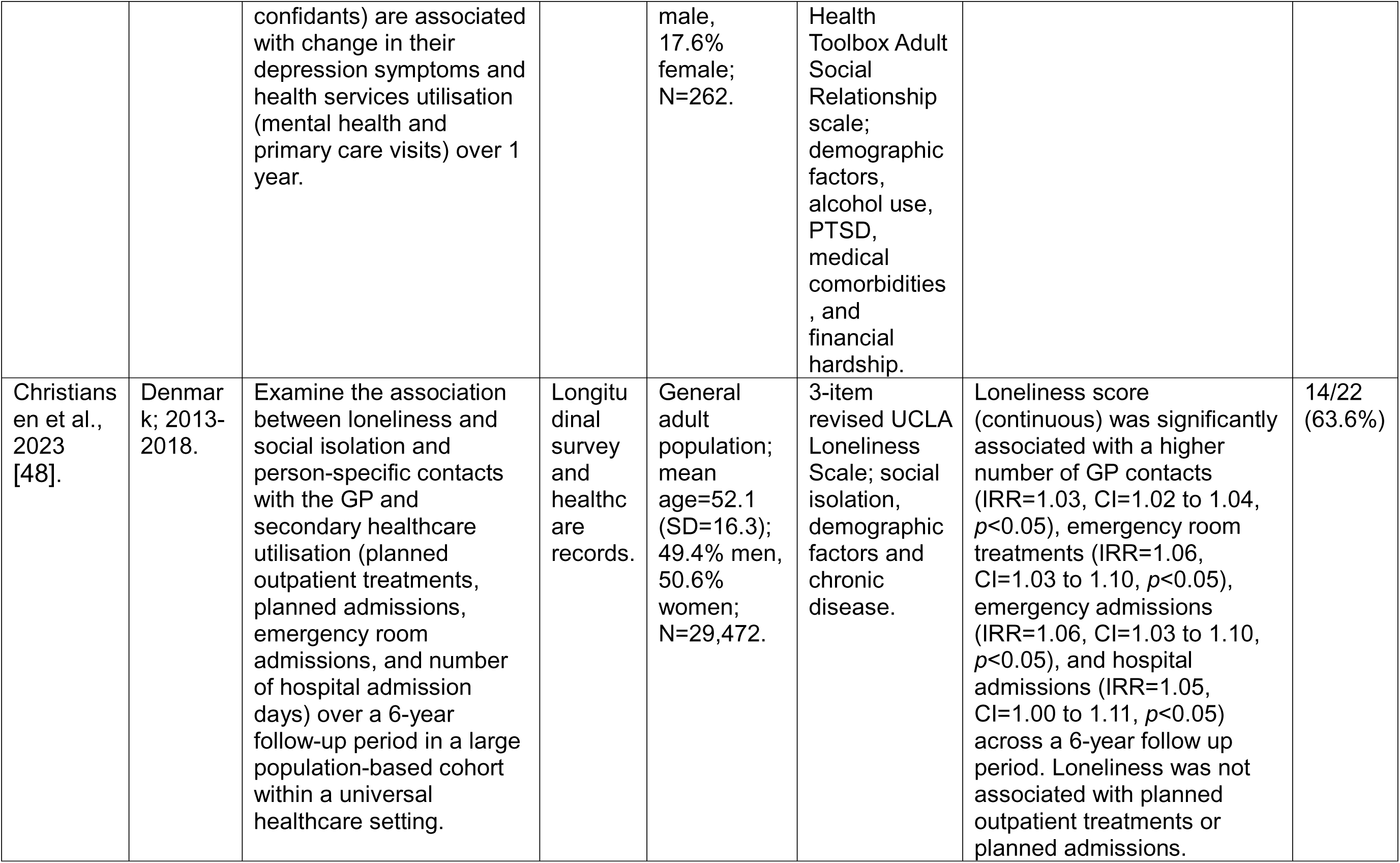
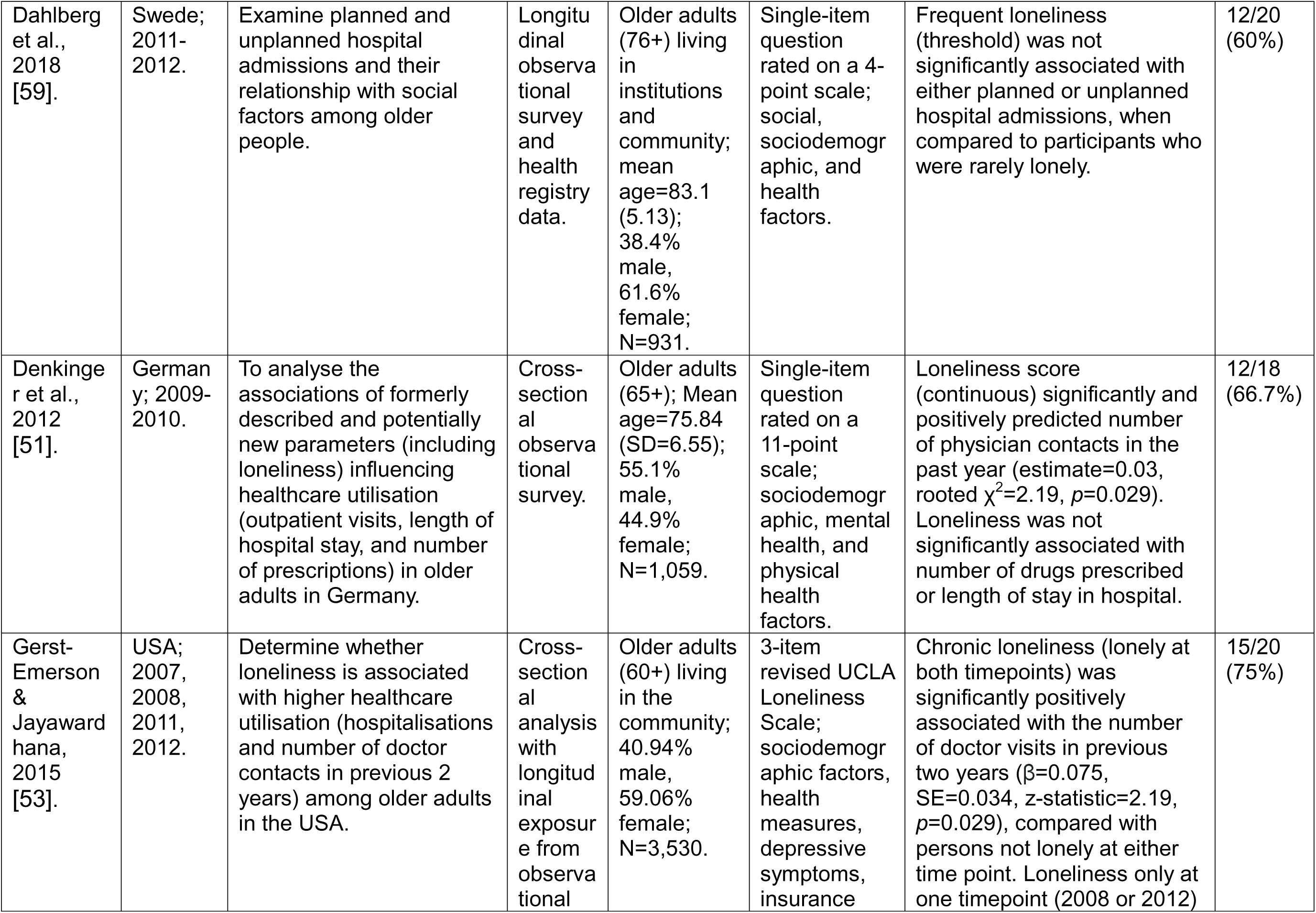
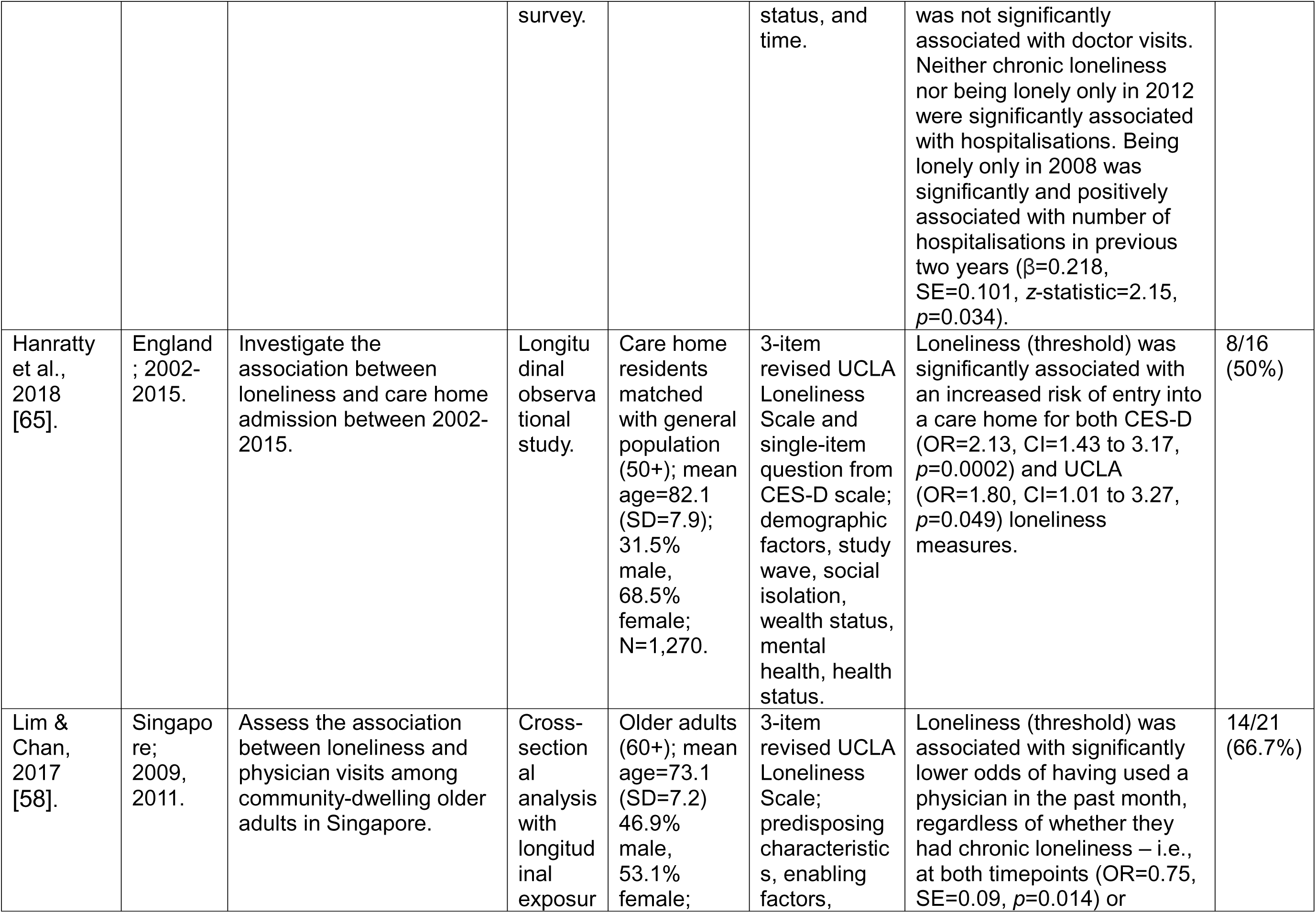
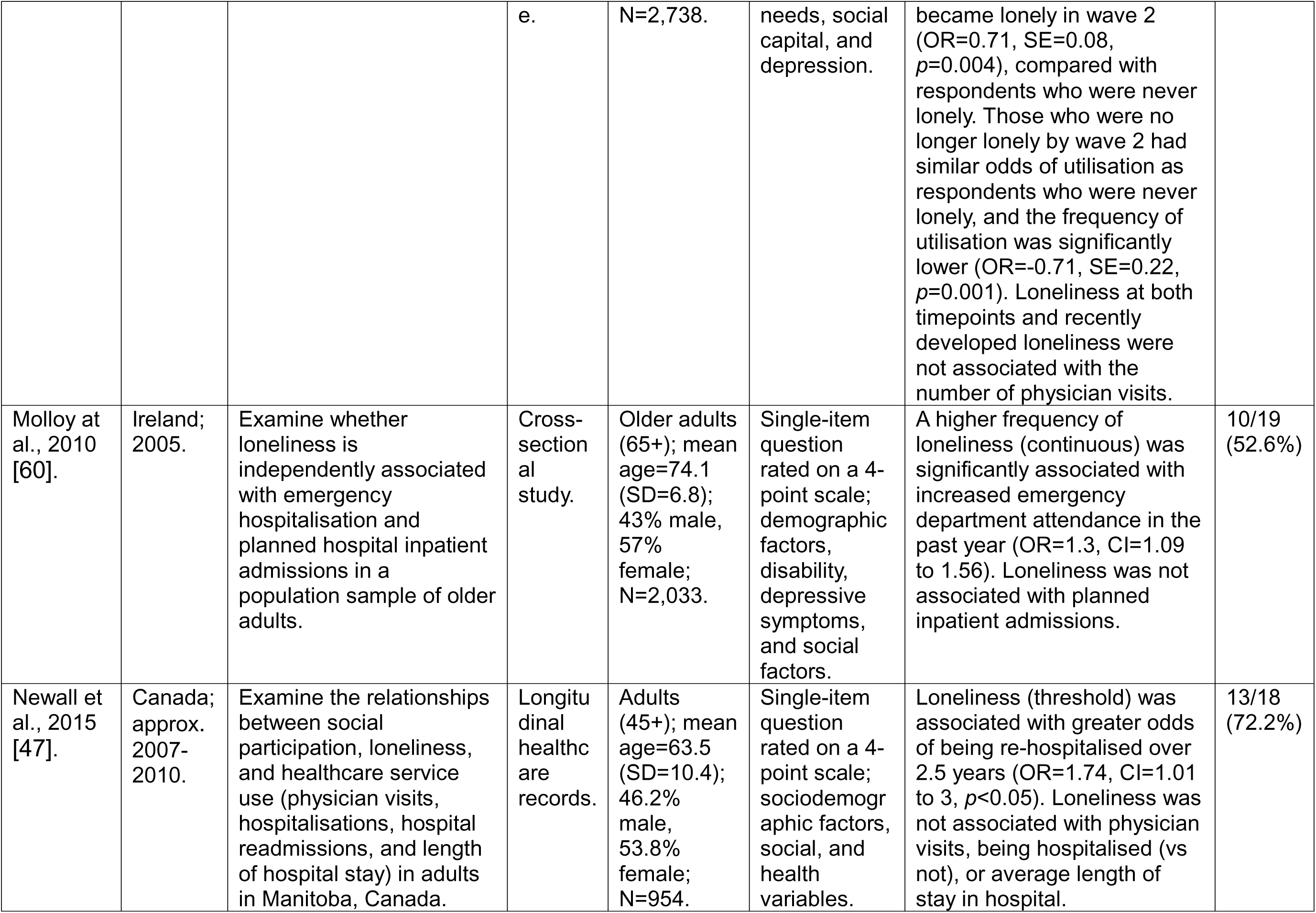
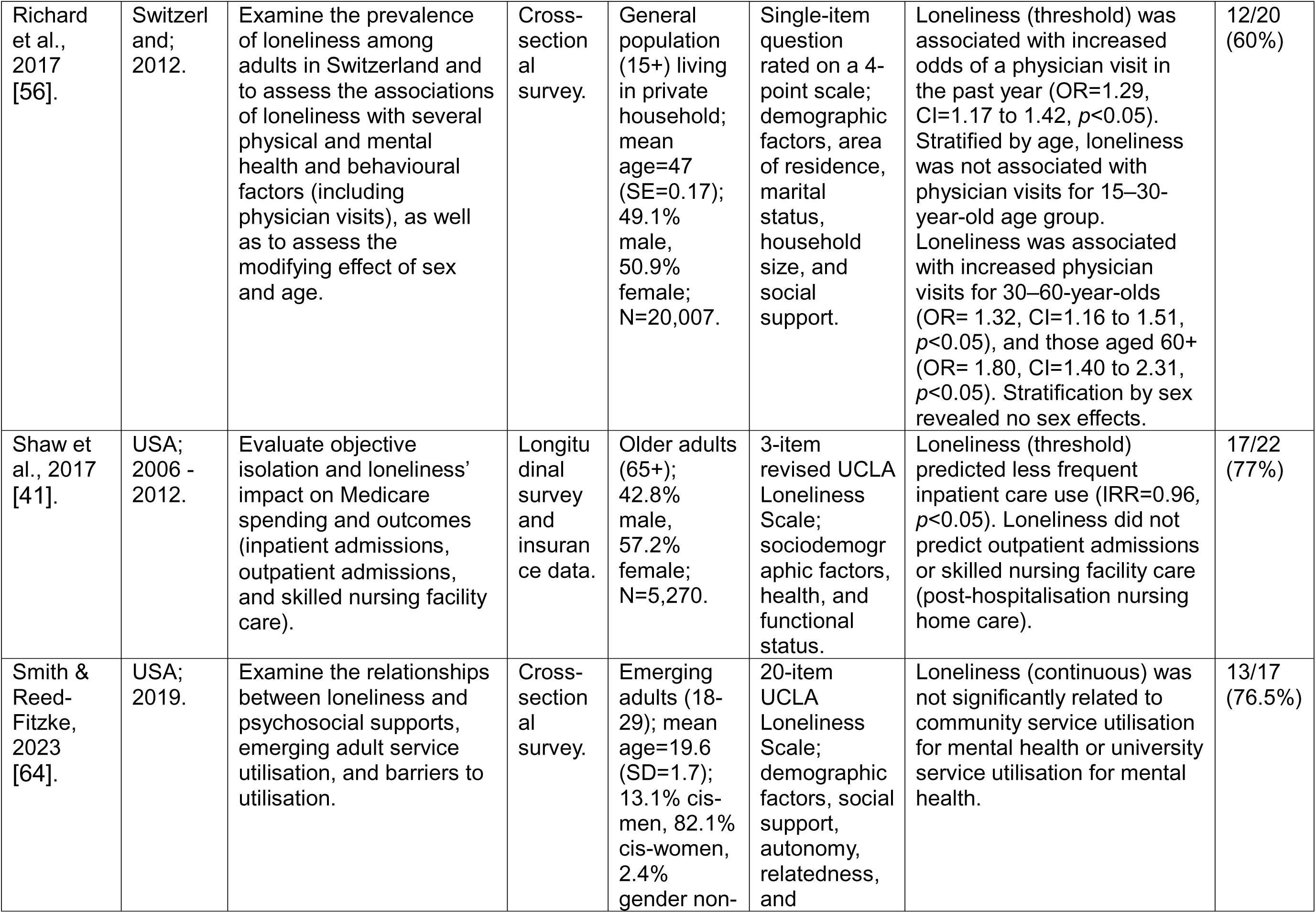
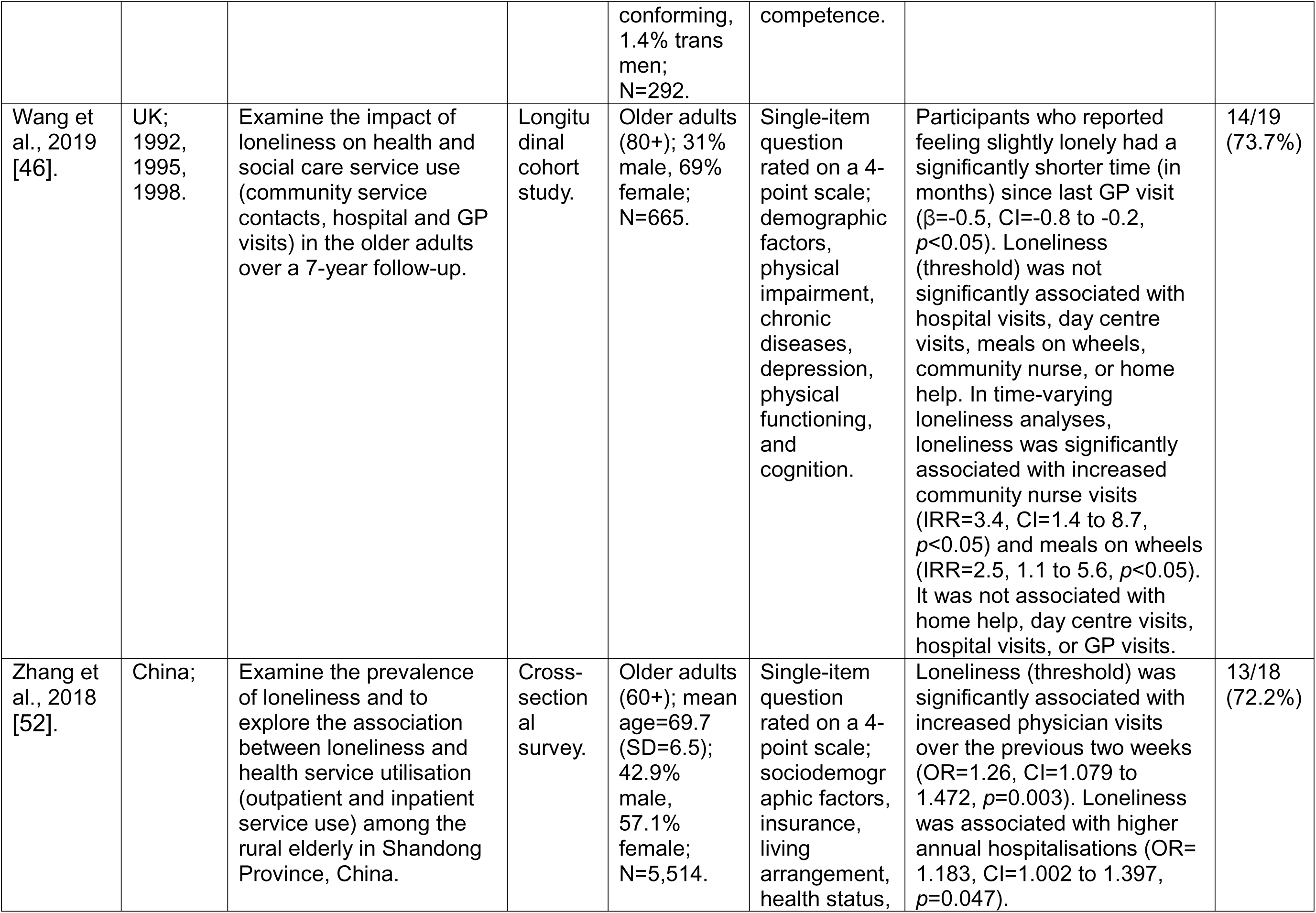
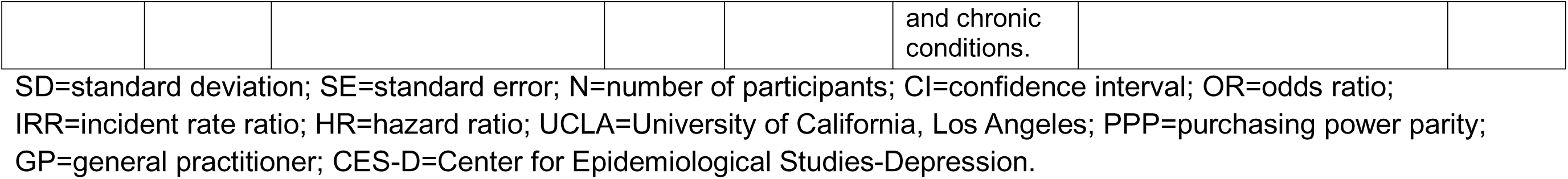
Summary of studies assessing the association between loneliness and healthcare use (n=22).

We identified 22 studies focused on healthcare use, three of which also included some measures of social care use. Of these, ten studies focused exclusively on older adult populations (aged 60 to 80+ years), six studies sampled middle-aged to older adults (aged 40 to 50+ years), two focused on young adults (aged 18 to 29 years), two sampled across the general population (aged 15+ to 18+ years and above), one studied military veterans (of all ages), and one focused on people with psychotic disorders (of all ages). The 22 studies in this category included fourteen cross-sectional studies and eight longitudinal studies. Eleven studies used a version of the UCLA Loneliness Scale to measure loneliness (ranging from the three-item version to the full 20-item version), nine used a single-item question, and the remaining two studies used the De Jong Gierveld loneliness scale and a loneliness subscale from the National Institute for Health Toolbox Adult Social Relationship scale, respectively. Ten of the studies were conducted in Northern and Western Europe (UK, Ireland, Switzerland, Denmark, Sweden, and Germany), nine in North America (Canada and USA), two in Asia (Singapore and China), and one in Australia.

None of the studies described any PPI or lived experience input to their research. Only one study, Wang et al. [46], mentioned PPI in their paper, acknowledging that PPI was not commonplace at the time of establishing the cohort study from which the data they analysed were drawn (early 1980s). Results are synthesised narratively below according to healthcare outcomes and service use.

### Loneliness and GP/physician visits

Thirteen studies in this category investigated the association between loneliness and GP or other physician visits. Two longitudinal studies judged to be of high methodological quality represented mixed findings: Newall et al. [47] found no significant evidence of an association between loneliness and having made one or more physician visits in adults (aged 45+) in a Canadian sample, while Wang et al. [46], considering a population of older adults in the UK (80+), showed that participants who reported feeling slightly lonely at baseline had a significantly shorter time (in months) since their last GP visit, compared to people who were not lonely at baseline (β=-0.5, CI=-0.8 to -0.2, *p*<0.05). They found no significant evidence of a shorter time interval since last GP visit for participants who reported feeling lonely *often*.

The remaining three longitudinal studies, which we found to be of moderate methodological quality, also reported a mixture of findings. When investigating the general adult Danish population, Christiansen et al. [48] found that loneliness was associated with a slight increase in the number of GP contacts (IRR=1.03, CI=1.02 to 1.04, *p*<0.05). A study investigating adults in Germany (aged 40+) did not find a significant association between loneliness and GP visits [49] and a study investigating military veterans in the USA did not observe a significant association between loneliness and primary care visits [50].

The other eight studies in this category were cross-sectional analyses and judged to be of moderate to high quality. Among older German adults (aged 60+), Denkinger et al. [51] identified a modest positive association between loneliness score and number of physician contacts in the past year (regression coefficient=0.03, rooted χ^2^=2.19, *p*=0.029). Zhang et al. [52] found an association between loneliness and increased physician visits among older adults in China (OR=1.26, CI=1.079 to 1.472, *p*=0.003), while Gerst-Emerson & Jayawardhana [53] observed a modest positive association between chronic loneliness (loneliness at two timepoints) and number of doctor visits in the USA (β=0.075, SE=0.034, *z*-statistic=2.19 *p*=0.029). In a Northern Irish sample, Burns et al. [54] found an association between loneliness scores and higher number of GP visits in middle aged to older adults (50+), although the magnitude of this effect estimate was small (IRR=1.03, CI=1.01 to 1.05, *p*=0.013). Burns et al. [55] found positive associations of a small magnitude between loneliness (IRR=1.03, CI=1.01 to 1.05, *p*=0.004) chronic loneliness (IRR=1.1, CI=1.01 to 1.19, *p*=0.028) and number of GP visits in middle aged to older adults in Ireland (aged 50+). They also found that women in their sample who reported loneliness at one time point and chronic loneliness (participants reporting loneliness at all three waves) had a slightly increased number of GP visits (IRR=1.05, CI=1.02 to 1.07, *p*<0.001; IRR=1.11, CI=1.01 to 1.23, *p*=0.03), while this did not apply to men. In a Swiss general population sample (aged 15 years and above), Richard et al. [56] found that loneliness was associated with increased odds of physician visits in the previous year (OR=1.29, CI=1.17 to 1.42, *p*<0.05). When stratified by age, this association was non-significant for 15–30-year-olds but was significant for those aged 30-60 years (OR= 1.32, CI=1.16 to 1.51, *p*<0.05) and those aged 60+ years (OR= 1.80, CI=1.40 to 2.31, *p*<0.05). Badcock et al. [57] found no significant association between loneliness and GP visits in those with psychotic disorder in an Australian sample.

In contrast to other results reported in this category, Lim and Chan [58] found that older Singaporean adults (aged 60+ years) who became lonely over time (OR=0.71, SE=0.08, *p*=0.004) and those who reported chronic loneliness – i.e., lonely at both timepoints (OR=0.75, SE=0.09, *p*=0.014) had lower odds of utilising a physician in the previous month, compared to those who were never lonely. Neither chronic loneliness nor recent loneliness were significantly associated with number physician visits in this sample.

Considered together, the findings from these studies provide little convincing evidence that loneliness is associated with an increase in number of GP visits or physician contacts, given that most of the studies that identified an association employed cross-sectional analyses and the majority of significant findings only showed minor changes in number of GP visits/physician contacts.

### Loneliness and hospital use/inpatient admissions

Twelve studies in this category considered the association between loneliness and hospitalisation/inpatient admissions. The three highest quality longitudinal studies considered several aspects of hospitalisation. Newall et al. [47] found that loneliness was not associated with risk of being hospitalised and was not associated with average length of stay in hospital, among Canadian adults (aged 45+). However, they did find that loneliness was associated with higher odds of re-hospitalisation over a period of 2.5 years (OR=1.74, CI=1.01 to 3, *p*<0.05). Wang et al. [46] did not observe a significant association between loneliness and number of hospital visits in adults in the UK aged over 80 years. In contrast, Shaw et al. [41] found that older adults in the USA (aged 65+) who reported loneliness reported slightly less frequent inpatient care admission (IRR=0.96, *p*<0.05).

Three longitudinal studies of moderate quality also examined the association between loneliness and hospitalisation. Christiansen et al. [48] observed a small but significant positive association between loneliness and number of days in hospital among Danish adults (IRR=1.05, CI=1.00 to 1.11, *p*<0.05), but not between loneliness and number of planned inpatient admissions. Dahlberg et al. [59] in a Swedish sample and Bock et al. [49] in a German sample found no significant association between loneliness and hospitalisation in older adults (aged 76+) and adults (aged 40+) respectively.

Two high quality cross-sectional studies conducted with data on older populations found some evidence that loneliness was associated with hospitalisations. In a Chinese sample, Zhang et al. [52] identified an association between loneliness and higher annual number of hospitalisations (OR=1.26, CI=1.079 to 1.472, *p*=0.003). In a sample from the USA, Gerst-Emerson & Jayawardhana [53] observed a modest association between loneliness and higher number of hospitalisations in the previous two years (β=0.218, SE=0.101, *z*-statistic=2.15, *p*=0.031). However, they only observed a significant association for people who were lonely in one specific point in time and not for chronic loneliness or loneliness at a different time points.

The remaining four studies in this category were cross-sectional and judged as moderate quality. These also presented mixed evidence. In an Australian sample, Badcock et al. [57] observed evidence of a positive association with loneliness and being a frequent hospital user, which included all health service use in the past 12 months (OR=1.8, *p*=0.03), and did not observe an association between loneliness and number of inpatient admissions in people with psychotic disorders. In older populations (aged 65+ years), Denkinger et al. [51] found no significant association between loneliness and length of stay in hospital (in a German sample) and Molloy et al. [60] did not find a significant association between loneliness and planned inpatient admissions (in an Irish sample). Barnes et al. [45] found that loneliness (and not social isolation) was not significantly associated with inpatient admissions among older adults in the USA.

In summary, we found little conclusive evidence that loneliness is associated with an increase in hospital and/or inpatient admissions in both longitudinal and cross-sectional studies.

### Loneliness and emergency department visits

Of the eight studies in this category that considered the association between loneliness and emergency department visits, only one was longitudinal. In this longitudinal study rated as moderate quality, Christiansen et al. [48] observed a significant association between loneliness and increased number of emergency room treatments/visits (IRR=1.06, CI=1.03 to 1.10, *p*<0.05) and number of emergency admissions (IRR=1.06, CI=1.03 to 1.10, *p*<0.05) in the Danish general adult population over a six year follow up.

The remaining seven studies in this category were all cross-sectional and of moderate quality. Molloy et al.[60] found that loneliness was associated with increased past-year emergency department attendance in older Irish adults (OR=1.3, CI=1.09 to 1.56), Chamberlain et al. [61] identified an association between loneliness and a higher number of emergency department visits in Canadian adults aged over 45 years (OR=1.13, CI=1.05 to 1.21), and Chamberlain et al. [62] found that risk of unplanned emergency department visits was higher for those reporting loneliness compared to those that did not report loneliness, in a sample of older Canadian adults in supported living (HR=1.10, CI=1.04 to 1.15). Chamberlain et al. [61] also found that loneliness was only significantly associated with increased emergency department visits among women (OR=1.15, CI=1.05 to 1.25), not men. Burns and colleagues [55] did not find a significant association between loneliness and emergency department visits in Irish adults (aged 50+). When stratified by sex they found that women who reported loneliness had an elevated risk of an emergency department visit (OR=1.08 CI=1.0 to 1.16, *p*=0.028), but not men. Burns et al. [54], Badcock et al. [57], and Barnes et al. [45], did not observe any significant associations between loneliness and emergency department visits in Northern Irish, Australian, or US samples, respectively.

In summary, we noted evidence from one longitudinal study that loneliness may be associated with increased emergency department visits, however findings from a range of cross-sectional studies were contradictory and therefore inconclusive.

### Loneliness and outpatient visits

Four studies examined the association between loneliness and outpatient care. Of these, three were longitudinal. In a high-quality longitudinal study of older adults in the USA, Shaw et al. [41] did not observe a significant association between loneliness and frequency of outpatient visits. The other two longitudinal studies were of moderate quality. Christiansen et al. [48] did not find a significant association between loneliness and planned outpatient treatments in the Danish adult population. Bock et al. [49] found no significant association between loneliness and specialist outpatient visits in adults (aged 40+) in Germany. One cross-sectional study judged to be of moderate quality [57] found no significant association between loneliness and outpatient visits in people with psychotic disorders in Australia.

In summary, there was therefore no evidence from these studies that loneliness is associated with frequency of outpatient visits.

### Loneliness and general mental health service use

Four studies considered mental health service use and its association with loneliness, of which only one was longitudinal. Chen et al. [50] examined the association between loneliness and mental health service use longitudinally (over 1 year) in veterans in the USA, finding no significant association in a study judged to be of moderate quality.

The remaining studies, all cross-sectional, were judged to be of moderate to high quality. Bessaha et al. [63] examined the association between loneliness and mental health service use in young adults (aged 18-29) in the USA, which was captured in terms of whether participants had received treatment and medication for their mental health, also finding no significant association. Smith & Reed-Fitzke [64] did not find a significant association between loneliness and community mental health service or university mental health service use, also within a young adult (aged 18 to 29) population in the USA. Badcock et al. [57] investigated the association between loneliness and home visits by a mental health professional in Australia, also finding no significant association.

Overall, there was no evidence from the few studies examining mental health service use that this was associated with loneliness.

### Loneliness and other measures of health and social care service use

Several studies considered other aspects of health and social care service use. Hanratty et al. [65] investigated the longitudinal association between loneliness and care home admission in English adults (aged 50+) in a study judged to be of moderate quality. They found that loneliness was significantly associated with increased odds of being admitted into a care home, based on two different measures of loneliness: the UCLA Loneliness Scale (OR=1.80, CI=1.01 to 3.27, *p*=0.049) and a single-item measure (OR=2.13, CI=1.43 to 3.17, *p*=0.0002).

Wang and colleagues [46] examined the longitudinal association between loneliness and different aspects of social care service use in a high-quality study of older adults (aged 80+) in the UK. They did not find an association between loneliness and day centre visits, meals on wheels, community nurse input, or home help when measuring loneliness at one time point. However, when measuring loneliness across three time points as a time-varying exposure, they identified an association between loneliness and increased community nurse visits (IRR=3.4, CI=1.4 to 8.7, *p*<0.05) and meals on wheels use (IRR=2.5, CI=1.1 to 5.6, *p*<0.05).

In another high-quality longitudinal study, Shaw et al. [41] found that loneliness did not predict changes in skilled nursing facility care (i.e., post-hospitalisation nursing home care) in older adults (aged 65+) in the USA. Finally, in a cross-sectional study, Denkinger et al. [51], found no association between loneliness and number of drugs prescribed in German older adults (aged 65+).

In summary, there is some longitudinal evidence that loneliness may be associated with specific aspects of social care use. However, outcomes were varied and findings inconsistent across studies.

### Economic evaluations of interventions to reduce loneliness

In total, we identified 18 economic evaluations from 14 publications, summarised in Table 3. Seven of the economic evaluations were cost-utility analyses (CUA), four were cost-consequence analyses (CCA), three were cost-effectiveness analyses (CEA), five were ROI analyses, and five were SROI analyses (note: several studies included more than one evaluation type). These studies were designed as follows: nine modelling studies, five impact evaluations of an intervention using pre-post study design, three randomised controlled trials (RCTs), and one service evaluation. Most studies used a validated measure to assess loneliness; seven used a version of the UCLA Loneliness Scale (ranging from the revised 3-item to the full 20-item version), six used a version of the De Jong Gierveld loneliness scale (including the full 11-item version and the revised 6-item version), and one used the 3-item Campaign to End Loneliness scale [66]. Of the remaining four studies, three identified loneliness through participant interviews and one was a modelling study that did not report a loneliness measure.

**Table 3.** Economic evaluation studies (n=18).

| *Lead author(s), year* | *Country; currency; costing year* | *Study aim* | *Study design; evaluation type* | *Sample description; sample size* | *Intervention description; comparator* | *Loneliness measure* | *Results* | *Quality score using Evers checklist [35]* |
| --- | --- | --- | --- | --- | --- | --- | --- | --- |
| Bauer et al., 2011 [74]. | UK; GBP; 2008-2009. | Estimate the cost-effectiveness of befriending interventions in terms of the reduction in depressive symptoms and the consequent decline in the use of health services by the recipient of the intervention. | Modelling study; ROI. | Lonely and isolated adults (50+). | One-on-one befriending intervention; no intervention. | None reported. | The ROI from an NHS perspective was £0.44 per £1 invested in the intervention over 1 year. Cost of befriending contact estimated at £85 (PPP=\$170.67). Intervention does not appear to be cost saving from public expenditure perspective. | 14/17 (82.4%) |
| Engel et al., 2021 [73]. | Australia; AUD; 2016. | Assess cost-effectiveness of a friendship intervention intended to reduce loneliness and depression in older adults. | Markov modelling study; CUA, ROI. | Older adults experiencing loneliness (55+); 100% female; N=163,229. | Friendship enrichment programme; usual care. | 11-item De Jong Gierveld loneliness scale | \$72.4 million (UI=-731 to 396; (PPP=\$63.84 million) of cost savings after 5 years due to reductions in healthcare treatment costs and productivity gains. Incremental cost-effectiveness ratio (ICER) was dominant: less costly and more effective than comparator. ROI of 2.87 (UI= -15.43 to 28.92) after 5 years. 7,889 quality adjusted life years (QALYs) gained (UI=-77,904 to 117,568) after 5 years. Some uncertainty on intervention effectiveness. | 17/19 (89.5%) |
| Engel et al., 2021 [73]. | Australia; AUD; 2016. | Assess cost-effectiveness of a computer and internet training programme intended to reduce loneliness and depression in older adults. | Markov modelling; CUA; ROI. | Older adults experiencing loneliness (65+); N=4,791. | Volunteer-led internet and computer training; usual care. | 6-item De Jong Gierveld loneliness scale. | \$4.7 million (UI=-38 to 9.9; PPP=\$4.14 million) in cost savings after 5 years. The ICER was dominant: less costly and more effective than comparator. ROI of 2.14 for volunteer led training (UI=-4.49 to 17.88) after 5 years. Resulted in 1,072 QALYs gained (UI=-3,939 to 8,569) after 5 years. Some uncertainty on intervention effectiveness. | 17/19 (89.5%) |
| Foster et al., 2021 [70]. | UK; GBP; 2020. | Assess the SROI of a link worker scheme to reduce loneliness in adults. | Pre-post impact evaluation with matched controls; SROI. | Adult service users of a social prescribing service; mean age=65.6 (SD=18.8) 33% male, 67% female; N=2,250 (up to 743 matched controls), N=4,010 for SROI analysis. | Social prescribed link workers providing up to 12 weeks of support to adults facing loneliness; no comparator, included matched control group. | 3-item revised UCLA Loneliness Scale. | Intervention resulted in a significant reduction in mean loneliness score of -1.8 (CI=-1.91 to -1.77, $p<0.001$). There was an estimated SROI of £3.42 per £1 invested in the service. Service was effective in reducing loneliness and returns exceeded investments. | 12/16 (75%) |
| Jones et al., 2015 [77]. | England; GBP; 2014. | Assess the feasibility of supporting volunteers in helping older people go online, assess the impact of | Pre-post impact evaluation; SROI. | Older adults (57+); 28.5% male, 71.5% female; N=144 | One-on-one and small groups covering basic computer use, how to get online | 6-item revised De Jong Gierveld loneliness scale. | Intervention resulted in a significant reduction in mean loneliness score of -0.58 ($p=0.004$). Cost of intervention was £708 (PPP=\$1,306.97) | 8/18 (44.4%) |
| | | using the Internet on contacts with others, loneliness, and mental health, and assess the perceived value to beneficiaries of going online. | | | and search the Internet, online shopping, email, Skype or FaceTime, and online news and entertainment; no comparator, participants were own controls. | | per person. Return estimated at £1,000 to £1,300 per person (PPP=\$1,846.01 to \$2,399.81). Therefore, intervention such as this may pay for itself in under one year. | |
| Jones et al., 2020 [71]. | Wales; GBP; 2018. | Assess SROI of a socially prescribed physical activity programme. | Pre-post impact evaluation; SROI. | Older adults (55+); mean age=72.4 (SD=8.3); 42.4% male, 57.6% female; N=66 participants (N=38 additional family members) | Health Precinct includes achievable exercise goals along with health support; no comparator, included matched control group. | 3-item Campaign to End Loneliness scale. | Value of benefits was £281,010 (PPP=\$488,198.73) and value of inputs was £55,389 (PPP=\$96,227.32). Social value of £5.07 for every £1 invested in the intervention. Sensitivity analysis yielded estimates of between £2.60 to £5.16 of social value. The social value generated by the intervention outweighed the cost of inputs required to deliver the programme. | 10/17 (58.8%) |
| Mallender et al., 2015 [80]. | England, data on intervention from USA; GBP; 2014. | Assess the cost and effects (in terms of raising awareness, identifying people at risk and/or improving mental wellbeing and/or independence) of a group-based singing intervention. | Modelling study; CCA. | Older adults; 22% male, 78% female; N=166 (N=90 intervention, N=76 control). | Group-based choir singing; usual activity (i.e. no choir singing) | 3-item revised UCLA Loneliness Scale. | There was no significant difference between groups on loneliness score. Cost per participant of intervention was £86 (PPP=\$158.76). There were 2.49 fewer doctor visits in intervention group, with costs averted of £92.13 (PPP=\$170.07) per participant. One fewer over the counter medication per year was associated with costs averted of £2.40 (PPP=\$4.43) per participant. Thus savings (£94.53; PPP=\$174.50) outweigh intervention costs. | 13/19 (68.4%) |
| Mallender et al., 2015 [80]. | England, data on intervention from the USA; GBP; 2013. | Assess the cost, effects (in terms of raising awareness, identifying people at risk and/or improving mental wellbeing and/or independence) | Modelling study; CCA, CUA. | Older adults; mean age intervention =71, mean age control=72; N=93 (N=48 intervention, N=45 control). | Group based internet and computer training intervention; no intervention. | 20-item UCLA Loneliness Scale. | There was no significant difference between groups on loneliness score at 5 months follow-up. Cost of intervention was £564 (PPP=\$1,054.46) per person. QALYs gained per person were 0.021 in | 15/19 (78.9%) |
| | | of a group Internet and computer training intervention. | | | | | intervention group. Incremental cost per QALY gained was £15,962 (PPP=\$29,842.59) and it was therefore deemed cost-effective. Cost savings associated with reduced adverse health conditions were £224 (PPP=\$418.79) per person, or savings of 40p per £1 spent on the intervention. | |
| Mallender et al., 2015 [80]. | England, data on intervention from the Netherlands; GBP; 2014. | Assess the cost, effects (in terms of raising awareness, identifying people at risk and/or improving mental wellbeing and/or independence) of a friendship programme. | Modelling study; CCA, CUA. | Older female adults; mean age=63; 100% female; N=115 (N=60 intervention, N=55 control). | Friendship programme for older people; no intervention. | 11-item De Jong Gierveld loneliness scale. | Loneliness significantly improved in the intervention group, but improvement was not significantly different to control group. QALYs gained per person in intervention group were 0.035 based on reductions in dis in a range of adverse health conditions associated with loneliness. Cost of the intervention was £77 (PPP=\$142.14) | 16/19 (84.2%) |
| | | | | | | | per person. Cost savings associated with reducing loneliness and health outcomes was £391 (PPP=\$721.79) per person, or a saving of £5.10 for every £1 spent. Intervention was dominant over control and was found to be both cost-effective and cost-saving. | |
| McDaid et al., 2017 [75]. | UK; GBP, 2015. | Assess impact of a signposting service for older people on service use (GP and GP nurse contacts, hospital presenting self-harm, and avoidance of psychological therapy for depression) and loneliness free life years, over five years. | Modelling study; ROI. | Older adults not in paid work (65+); N=100,000. | Signposting service for people not in paid work; no intervention. | Self-identified as being lonely. | Over five years, from a societal perspective there was a ROI of at least £1.26 from every £1 invested in this service. Intervention resulted in 1,313 extra loneliness-free years. | 13/18 (72.2%) |
| McDaid et al., 2021 [72]. | UK; GBP; 2016. | Examine the impact of participation in Reconnections | Impact evaluation; CEA. | Older adults (50+), mean age=74.9; 26.4% men, | Personalised support and community | 4-item revised UCLA Loneliness | Loneliness scores improved significantly in the intervention group | 15/18 (83.3%) |
| | | on the use of health and social care services, as well as capture qualitative insights into the experience of participating in Reconnections. | | 73.6%;<br>N=121 | response to loneliness;<br>no comparator. | Scale. | ($p<0.0001$), maintained at 18-month follow up. Intervention had a mean cost of £752 (PPP=\$1388.20) per person. Little difference in health service costs pre and post intervention. Outpatient costs were significantly lower post intervention ($p=0.034$). Loneliness improvement was associated with 32% lower A&E costs ($p=0.02$). | |
| McDaid et al., 2021 [72]. | UK; GBP; 2019. | Look at the potential impact on health, social care and informal care costs over a five-year period of the Reconnections service and net costs averted as a result of reducing levels of loneliness. | Modelling study; ROI, cost-consequences analysis. | Older adults (50+)<br>N=500. | Personalised support and community response to loneliness;<br>no comparator. | 4-item revised UCLA Loneliness Scale. | Intervention had a mean cost of £752 (PPP=\$1,279.41) per person. Net positive gain of £41,000 (PPP=\$69754.86) over 5 years. Total ROI of £1.11 for every £1 invested. Results in an extra 486 loneliness-free years gained. | 13/18<br>(72.2%) |
| McDaid & Park., 2021 [81]. | England; GBP; 2019. | Model synthesises short term costs and experience in tackling loneliness in England with information on the potential health and social care benefits that may be gained through a reduction in loneliness over a 5-year time period. | Markov modelling study; CEA. | Older people with moderate or severe loneliness (65+); N=1,200. | Local signposting service to help older people make new social connections in their community; no intervention. | 4-item revised UCLA Loneliness Scale. | Mean 0.45 additional loneliness-free years would be gained from the intervention, over 5 years. Cost per participant in intervention group was £7,131 (PPP=\$12,132.24) compared to £6,783 (PPP=\$11,540.18) in usual care. Incremental cost per loneliness-free year gained was £768 (PPP=\$1,306.63). Intervention was cost saving in 3.5% of iterations. Intervention was likely to be cost-effective but unlikely to be cost saving. | 17/19 (89.5%) |
| Mountain et al., 2017 [67]. | UK; GBP; 2013-2014. | Test whether an intervention modelled on Lifestyle Redesign and adapted for a UK population (Lifestyle Matters) could also demonstrate clinical and cost- | RCT; CUA. | Older adults (65+); mean age=72.1; 31.9% male, 68.1% female; N=262 (N=136 intervention, N=126 control). | Manualised *Lifestyle Matters* intervention, an occupation-based lifestyle intervention; usual care. | 11-item De Jong Gierveld loneliness scale. | No evidence of intervention effect at 6 months on any outcomes. At 24 months, loneliness was significantly improved in the intervention compared to the control group (mean difference=-0.7, CI=-1.4 to -0.1, $p=0.026$). | 17/19 (89.5%) |
| | | effectiveness. | | | | | The ICER was £7,621 (PPP=\$14,068.44). Probability of 30% that intervention would be cost-effective. Little evidence of clinical or cost-effectiveness. | |
| Simpson et al., 2014 [78]. | UK; GBP; 2010. | Investigate the effect of peer support on feelings of hope and loneliness, quality of life, and service use in mental health patients following discharge from hospital. Evaluate the economic consequences of the intervention, with follow-up at one and three-months post-discharge. | Pilot RCT; CEA and CUA. | People with mental illness diagnosis; mean age intervention =34.13 (SD=10.27), and control =23.36 (SD=10.15); intervention 69.6% male, 30.4% female, control 87% male, 13% female; N=46 (23 intervention, 23 control). | Peer support for patients recently discharged from inpatient care; care as usual. | 20-item UCLA Loneliness Scale. | There were no significant differences between groups on loneliness or other outcomes. Total cost per case for intervention was £2,154 (\$4,263.99) and for control was £1,922 (\$3,804.73). Mean difference between costs was not significant. Peer support intervention did not show cost-effectiveness. | 17/18 (94.4%) |
| Social Value Lab; 2011 [69]. | UK; GBP; 2010-2011. | Assess the social value of a Craft Café for older people. | Impact evaluation; SROI. | Older people (50-90); N=72. | Craft Café to enable skill acquisition, | Reporting of reduced loneliness via | The total benefits for the programme were valued at £696,569 (PPP=\$1,350,217.41 | 13/19 (68.4%) |
| | | | | | improve social networks, and connections with community; no comparator. | baseline and tracking interviews with participants and family, and tutor feedback. | ). Programme cost was valued at £84,219 (PPP=\$163,248.67). Social value of £8.27 for every £1 invested (sensitivity analyses ranged from £4.86 to £9.57 of social value). | |
| Weiss et al., 2020 [79]. | Netherlands; EUR; 2016. | Evaluate the cost-effectiveness of a positive psychology intervention. | RCT; CUA. | Adults experiencing loneliness, experiencing mental and/or physical health problems, and with low socio-economic status; median age=60 (IQR=48.3-68.0); 30.6% male, 69.4% female; N=108 (N=58 | Happiness Route positive psychology programme; active control of customised care from counsellors. | 11-item De Jong Gierveld loneliness scale. | No significant difference between groups on loneliness, though both groups had significantly improved loneliness scores ($p<0.05$). Intervention group gained 0.07 (CI=0.05 to 0.11) less QALYs on average compared to control group. Cost of the intervention was €1,090 (PPP=\$1705.08) higher in control group. Cost saved per lost QALY was €161,953 (PPP=\$253,341.64). Intervention had 84% likelihood of cost-effectiveness, though accrued less | 14/18 (77.8%) |
|  |  |  |  | intervention, N=50 control). |  |  | QALYs. |  |
| Willis et al., 2018 [68]. | UK; GBP; 2014-2015. | Quantify the social value of peer support groups for people with dementia and their carers. | Service evaluation; SROI. | People with dementia and their carers; group A: N=23, group B: N=5, group C: N=9. | Peer support groups for people with dementia and their carers; no comparator. | Loneliness was self-identified in through qualitative interviews. Financial proxy for loneliness was cost of treating someone for depression in NHS. | Three groups created social value ranging from £1.17 to £5.18 for every £1 of investment, dependent on the design and structure of each group. Key outcomes for people with dementia included reduction in loneliness and isolation. | 13/16 (81.3%) |

SD=standard deviation; N=number of participants; CI=confidence interval; UI=uncertainty interval; OR=odds ratio; IRR=incident rate ratio; UCLA=University of California, Los Angeles; PPP=purchasing power parity; GP=general practitioner; A&E=accident and emergency; ROI=return on investment; SROI=social return on investment; CEA=cost-effectiveness analysis; CUA=cost-utility analysis; CCA=cost-consequence analysis; QALY=quality adjusted life years.

The majority of studies examined populations aged 50 years and older. Seven studies specified older adult populations (three studies with participants aged 65+ and four studies without a specified age range) and seven evaluations were with middle-aged and older adult participants (aged 50+ to 57+). Of the remaining evaluations, one analysed data from a general adult population, one for people with a mental illness diagnosis, one for adults who reported mental and/or physical health problems and low socio-economic status, and one for people with dementia. A total of 15 studies were conducted or modelled in the UK, whilst two were set in Australia, and one in the Netherlands.

A total of 12 out of the 18 evaluations did not mention any PPI or lived experience involvement. Only one study mentioned having consulted one individual with lived experience to review their study protocol and supporting documents [67]. As part of their SROI analyses, Willis and colleagues [68], the Social Value Lab [69], Foster et al. [70], and Jones et al. [71] consulted a range of stakeholders, including participants of each respective intervention. The study by McDaid and colleagues [72] also included perspectives from participants in the study intervention when reporting their CEA. However, none of these analyses reported co-producing or consulting with individuals with lived experience in the design or running of their studies.

### Return on investment studies

The five ROI analyses were all from high quality modelling studies. Engel et al. [73] conducted two ROI analyses; one was an assessment of a friendship enrichment programme and one was an assessment of a computer and internet training programme, both intended to reduce loneliness and depression in older adults in Australia. The friendship enrichment programme was modelled in a large sample of women (N=163,299) aged over 55, and when compared to usual care had a ROI ratio of $2.87 for every $1 invested (AUD; UI=-15.43 to 28.92) after 5 years. In related analyses, the intervention was found to have cost savings of $72.4 million (AUD; UI=-731 to 396; PPP=$63.84 million) after 5 years due to reductions in healthcare treatment costs and productivity gains, with 7,889 QALYs gained (UI=-77,904 to 117,568) and a dominant incremental cost-effectiveness ratio (ICER). However, strength of evidence of the effectiveness of the intervention in reducing loneliness was poor. Similarly, the computer and internet training programme was modelled in a sample of adults aged over 65 (N=4,791), and, when compared to usual care, had a ROI ratio of $2.14 for every $1 invested (AUD; UI=-4.49 to 17.88) after 5 years. Further analyses demonstrated cost savings of $4.7 million (AUD; UI=-38M to 9.9M; PPP=$4.14 million), 1,072 QALYs gained (UI=-3,939 to 8,569), and a dominant ICER, however there was also uncertainty over the effectiveness of this intervention in reducing loneliness.

Bauer and colleagues [74] examined the cost-effectiveness of a one-on-one befriending intervention for adults in the UK aged over 50, compared to no intervention. They found a ROI of £0.44 per £1 invested in the intervention, from an NHS perspective. Thus, the intervention did not appear to be cost saving from a public expenditure perspective. McDaid et al. [75] modelled the impact of a signposting service for people over 65 not in paid work on a sample of 100,000 people, also in the UK. Over 5 years, taking a societal perspective, they found that the intervention had a ROI of £1.26 from every £1 invested and resulted in 1,313 additional loneliness-free years. McDaid et al. [72] also modelled the impact of a personalised support and community response programme for loneliness in UK adults over 50 (N=500) over 5 years. They observed a ROI of £1.11 for every £1 invested in the intervention and an extra 486 loneliness-free years gained.

In summary, this evidence demonstrates a positive ROI value for several loneliness interventions, though their effectiveness in reducing loneliness was not consistently established.

### Social return on investment studies

All five SROI studies were carried out within the UK. Unlike ROI studies, which compare the value of monetary costs averted with the additional costs of intervention, SROI studies additionally place a monetary value on benefits that are difficult to value monetarily. This might be done after consultation with relevant stakeholders on why and how they believe an action will work, as well as on the magnitude of these effects [76]. The SROI study that was rated as the highest quality was conducted by Willis et al. [68].

This examined the social value of three peer support groups for people with dementia and their carers. The three groups were estimated to create social values ranging from £1.17 to £5.18 for every £1 invested. The social value generated depended on the design and structure of each group, and key outcomes included a reduction in loneliness. The group that had an SROI of £5.18 was conducted weekly, included carers in the group, had approximately 23 participants in each session, focused on a variety of group activities, and had 10 volunteer staff involved as well as paid staff and a group facilitator. The other two groups, which had SROI values of £1.71 and £1.17 respectively, occurred less frequently (fortnightly and monthly, respectively), had fewer participants (5 and 9, respectively), fewer volunteers (zero and two, respectively) and were focused on group activities related to memory and reminiscence. The other high quality SROI study was conducted by Foster and colleagues [70] and assessed the social value of a link worker scheme to reduce loneliness in adult service users of a social prescribing service (N=4,010). They found an estimated SROI value of £3.42 per £1 invested in the service and the intervention significantly reduced mean loneliness score on the 3-item UCLA Loneliness Scale by 1.8 (CI=-1.91 to -1.77, *p*=<0.001). The authors concluded that the service was effective in reducing loneliness and that the social value returned exceeded the investment.

In a moderate quality study, the Social Value Lab[69] conducted SROI analysis of a Craft Café in a small sample of older adults aged 50-90 (N=72). The intervention was designed to enable skills acquisition and improve social and community connections. There was an estimated SROI value of £8.27 for every £1 invested. Sensitivity analyses produced estimates ranging from £4.86 to £9.57 and participants discussed feeling that their loneliness had reduced in qualitative interviews, though this was not assessed quantitatively. Two further SROI analyses in this category were of moderate quality. Jones et al.[71] assessed the SROI of a social prescribing physical activity programme for a small group of adults over 55+ (N=66). They observed a SROI value of £5.07 generated for every £1 invested in the programme, with sensitivity analyses identifying estimates ranging from £2.60 to £5.16, suggesting that the intervention outweighed the input costs. Finally, Jones et al.[77] conducted a SROI of an intervention supporting volunteers to help people aged 57+ access and use the internet (N=144). The intervention resulted in a significant reduction in mean De Jong Gierveld loneliness score of 0.58 (*p*=0.004). The cost of the intervention was £708 (PPP=$1,306.97) per person with a SROI value estimated at £1,000 to £1,300 (PPP=$1,846.01 to $2,399.81) per person. Therefore, the authors concluded that the invention would theoretically pay for itself in under one year.

In summary, the loneliness interventions evaluted in this category demonstrated positive SROI values, suggesting that they can generate social value.

### Cost consequence, cost-utility, and cost-effectiveness analyses

The eight evaluations in this category were either a CCA, CUA, or CEA, or a combination of these approaches. Three of these studies were incorporated into RCTs. The highest quality study was a pilot RCT conducted in the UK by Simpson and colleagues [78] to examine the effect of peer support for a small sample of patients recently discharged from inpatient care, with care as usual as the comparator (N=46). Their CEA and CUA found no significant mean difference between costs for the two groups and no significant difference between groups on loneliness outcomes at three months post-discharge. Mountain et al. [67] conducted a high-quality RCT in the UK to examine an occupation-based lifestyle intervention for older adults aged 65+, compared to usual care (N=262), also incorporating a CUA. They found loneliness, measured using the 11-item De Jong Gierveld scale, to be significantly lower in the intervention group at 24 months post-intervention (mean difference=-0.7, CI=-1.4 to -0.1, *p*=0.026) but not at 6 months. The ICER was £7,621 (PPP=$14,068.44). They concluded that the probability of the intervention being cost-effective was only 30%, and that there was little convincing evidence of clinical or cost-effectiveness of the intervention.

The third RCT in this category was conducted by Weiss and colleagues [79] in the Netherlands, to compare a positive psychology intervention for adults experiencing loneliness, experiencing mental and/or physical ill health, and with low socioeconomic status (N=108) to an active control of customised care from counsellors. Both groups significantly improved on loneliness, but with no significant difference between them. Although the intervention group gained 0.07 less QALYs on average than the control group (CI=0.05 to 0.11), the intervention cost was €1,090 higher in the control group and the cost saved per lost QALY was €161,953 (PPP=$253,341.64). The authors concluded that the intervention had an 84% chance of being cost-effective but may not be considered a good investment by some policy makers as it also accrued less QALYs than the control group.

McDaid et al.[72] conducted an impact evaluation and CEA of a personalised support and community response to loneliness, named *Reconnections,* which had been delivered to older adults in the UK. In an analysis judged to be high-quality, using data from a sample of 121 participants they found that loneliness scores on the 4-item UCLA Loneliness Scale improved significantly in the intervention group (*p*<0.0001), and that this was maintained at the 18-month follow-up. The intervention had a mean cost of £752 (PPP=$1388.20) per person, and there was no significant difference between health service costs pre- and post-intervention. Outpatient costs were found to be significantly lower in the intervention group (*p*=0.034) and the improvement in loneliness score was associated with 32% lower A&E costs (*p*=0.02).

The four remaining studies in this category were modelling studies, all using UK data. Mallender and colleagues [80] conducted three of these studies, while McDaid & Park [81] carried out the fourth. The highest quality of these was by McDaid and Park [81], who assessed a local signposting service to help older people aged 65+ years who reported moderate or severe loneliness to make new social connections in their community, compared to usual care (N=1,200). This CEA identified a mean additional 0.45 loneliness-free years gained from the intervention over 5 years. The cost per participant in the intervention group was £7,131 (PPP=$12,132.24) compared to £6,783 (PPP=$11,540.18) in the usual care group. The incremental cost per loneliness-free year gained over 5 years was £768, and the intervention was cost saving in 3.5% of iterations. It was concluded that the intervention was likely to be cost-effective, but unlikely to be cost saving.

In a modelling study also judged to be high-quality, Mallender et al. [80] assessed the cost-effectiveness of a friendship programme for older female adults, compared to a group receiving no intervention (N=115). While the economic analyses were modelled in a UK population, the original intervention was conducted in the Netherlands. The CCA revealed that loneliness significantly improved in the intervention group over 9 months, but that there were no significantly differences to the control group. The intervention was estimated to cost £77 (PPP=$142.14) per person. In a CUA, 0.035 QALYs were gained per person based on reductions in disutility across a range of adverse health conditions associated with loneliness. It was concluded that the cost savings associated with reducing loneliness and associated health outcomes were £391 (PPP=$721.79) per person, or savings of £5.10 per £1 spent and a net saving of £314 (PPP=$579.65) per person. Therefore, the intervention was dominant over the control – i.e. both more effective and with lower costs.

Mallender et al. [80] modelled the cost-effectiveness of a group internet and computer training intervention for older adults in a high-quality study, with no intervention as the comparator (N=93). The study was modelled on a UK population, while the original intervention was from the USA. The CCA showed no significant impact on loneliness in the intervention group, after a 5-month follow-up. The intervention was estimated to cost £564 (PPP=$1,054.46) per person. Their CUA revealed 0.021 QALYs gained in the intervention group. The incremental cost per QALY gained was £15,962 (PPP=$29,842.59), so the intervention was deemed cost-effective. Cost savings associated with reduced loneliness were £224 (PPP=$418.79) per person, or savings of 40p per £1 spent on the intervention.

Another modelling study, also by Mallender et al. [80] was a moderate to high quality analysis of the cost-effectiveness of a group choir singing intervention in older adults, compared to no intervention (N=166) and followed up after 12 months. This analysis was modelled in a UK population. A CCA was conducted and showed no significant difference between groups on loneliness score at the 12-month follow-up. The cost of the intervention was £86 (PPP=$158.76) per participant. Costs averted per participant were £92.13 (PPP=$170.07) for 2.49 fewer doctor visits and £2.40 (PPP=$4.43) averted for fewer medications in the intervention group. Total savings therefore outweighed intervention costs.

Overall, findings demonstrated that several different types of loneliness interventions had the potential to be cost-effective, though general effectiveness in reducing loneliness was limited.

## Discussion

### Summary of findings

Our review synthesised evidence from 39 studies that examined the healthcare impact of loneliness, and the cost-effectiveness of loneliness interventions. We identified five studies that estimated either the healthcare cost or expenditure associated with loneliness; two of these studies identified a significant association between loneliness and lower healthcare expenditure, while one study reported higher expenditure for lonely individuals. In terms of cost, one study modelling the healthcare cost of loneliness for the public sector reported excess healthcare costs in the medium term, whereas another study found no statistically significant difference in direct healthcare costs between lonely and non-lonely individuals. The inconsistency of this evidence, represented by a small number of studies from a variety of contexts and using differing methods, presents a challenge to drawing robust conclusions about the impact of loneliness on healthcare costs and expenditure. Furthermore, only two studies directly assessed healthcare costs associated with loneliness, while the remainder focused on healthcare expenditure. This highlights a clear lack of research on the true cost of loneliness, posing a considerable challenge for assessing its economic impact. It is also important to note that four of these studies focused on older adult populations, and the sample of adults in the remaining study had a mean age of 59.3 years. These studies also primarily focused on healthcare costs and expenditure and generally did not consider wider social care costs, which may have particular and increasing relevance for older adult populations [82].

Across the 22 studies examining the association between loneliness and healthcare use, there was little consistent evidence that loneliness was associated with GP/physician contacts, hospital/inpatient admissions, outpatient visits, or mental health service use. The majority of research into loneliness and healthcare use specifically considered GP/physician visits, inpatient/hospital admissions, and emergency department visits, with less evidence for studies examining outpatient visits, mental health service use, and social care use. Relatedly, most of the high-quality longitudinal evidence related to GP/physician visits and inpatient/hospital admissions. Even so, much of the high-quality longitudinal evidence within different types of healthcare use is inconsistent, and in some instances, findings were contradictory. Therefore, an overall robust conclusion cannot be drawn about the association between loneliness and healthcare use from the findings identified in this study.

We identified 18 economic evaluations from 14 studies in this review. The findings from these studies suggest that loneliness interventions can be cost-effective, can generate social value, and that reducing loneliness may improve health related quality of life. However, few interventions were identified as potentially cost saving and their effectiveness in reducing loneliness was not robustly established. While many of the studies identified were rated as high quality, only three studies were RCTs. Further, only one of these studies concluded that their intervention, a positive psychology programme, was likely to be cost-effective, also noting that it may not be considered a good investment by policy makers as it also accrued fewer QALYs than the control condition [67]. This highlights the importance of conducting high quality evaluations of interventions alongside thorough economic evaluations, before robust conclusions can be drawn about intervention efficacy and cost-effectiveness. The lack of such studies is a notable limitation of research on this topic.

It should also be noted that the economic evaluations we identified related to a wide range of different intervention types, including several befriending interventions, peer support interventions, computer training programmes, signposting, social prescribing, and others. The studies also employed a variety of evaluation methodologies. This makes comparisons across different interventions and studies difficult. Furthermore, many of the sample sizes were small, so results should be interpreted with caution. All of the interventions were aimed at adult populations, and the majority focused on older adults. Therefore, findings are likely to be most relevant for this group and may not be applicable to younger populations. It is also important to note that 15 of the 18 economic evaluations were conducted or modelled in UK populations, so findings are likely to be most relevant to a UK context.

### Strengths and limitations

A notable strength of the current review is our inclusion of studies that measure the healthcare cost/expenditure of loneliness, the impact of loneliness on healthcare use as well as studies that examine the cost-effectiveness of loneliness interventions. As such, this review considers the healthcare impact of loneliness from several key perspectives, representing a thorough evaluation of existing research and building on previous reviews on this important topic.

We addressed important limitations of the review conducted by Mihalopoulos et al. [29] by including studies that included both mental and physical health outcomes and studies that included loneliness as a secondary outcome (provided loneliness was a main focus of the paper). We also addressed important limitations of the review conducted by Smith et al. [30], by including studies with any population, including those who were not older adults and those beyond the general population. We also did not limit our search to studies published in English, addressing a limitation of both previous reviews.

It is also important to note several limitations of the current review. The studies included in this review are predominantly focused on older populations. This is despite loneliness being at least as common among adolescents/young adults as in older adults [5–7]. For instance, research in the UK has previously found that young people aged 16 to 24 years report a higher prevalence of frequent loneliness than any other age group in the population [83]. This highlights a key research gap, as the economic impact of loneliness in groups other than older adults has not been well examined. This is especially true for economic evaluations of loneliness interventions, none of which were conducted with younger populations. Given this, the findings in this review are likely to be most relevant for older age groups.

Furthermore, the studies included in this review were from a small number of countries, predominantly within the UK and the USA. Therefore, findings are most likely to reflect these contexts and may not represent other countries. Additionally, many of the included studies employed cross-sectional designs, limiting understanding of the temporality of identified associations and of the economic impact of loneliness over time. Both the cross-sectional and longitudinal studies may also include participants who were lonely prior to the study period. This may mean that associated costs and/or service use may already have been incurred before the study period, and newer impacts of loneliness may not be reflected. Relatedly, few of the longitudinal or intervention studies had follow-up periods longer than ten years. It is possible, therefore, that the impact of loneliness on health and social care use, and the outcomes of loneliness interventions, may occur over longer time periods than were captured by these studies.

Our focus in this review was on studies relating to health and social care use. This means that it is likely we have missed studies that consider economic impacts beyond health and social care, such as economic productivity in workplaces, informal care, and volunteering. These are important considerations on this topic and should be examined in future research. The general focus on academic literature may also mean that we missed loneliness interventions offered by charities and voluntary sector organisations, as these sources may have fewer resources to undertake economic evaluations. However, we attempted to mitigate this through thoroughly searching two sources of grey literature and backward citation searching of relevant reviews.

As evidence was generally mixed and imprecise, we opted to grade the overall certainty of evidence as low and to weight the strength of our recommendations accordingly, instead of using the Grading of Recommendations Assessment, Development, and Evaluation (GRADE) guidelines[84]. This represents a deviation from our study protocol and is a limitation of this review. Finally, we only included studies that examined loneliness and did not consider related concepts such as social isolation, social exclusion, and perceived social support. Future studies should aim to assess the health and social care impact of these related domains and whether they differ to the impact of loneliness.

### Implications for future research

This is an important and topical research area, likely to gain further relevance in coming years due to current economic pressures and an increase in loneliness-related research [27]. As such, continued research into the health and social care impact of loneliness is warranted to determine how best to direct investment to address this problem. Specifically, our review identified a lack of studies looking at the healthcare cost of loneliness, which are essential to understanding its economic impact on the healthcare system. High-quality longitudinal studies should also be a priority, with extended follow-up periods to identify whether there are long term impacts of loneliness on health and social care use. An additional important consideration is to explore the association between loneliness and help-seeking behaviour and whether this has an impact on healthcare use, and associated costs, over time. Continued focus on identifying loneliness interventions that are both effective and cost-effective is also warranted. More RCTs of loneliness interventions with long follow-up periods, detailed cost analyses, and qualitative components to understand participants’ experiences could improve understanding of the acceptability, effectiveness and cost-effectiveness of loneliness interventions over time.

One finding of potential note derives from two cross-sectional analyses that showed an association between loneliness and emergency department visits in women but not in men. This highlights a possible future research avenue to examine sex differences in the association between loneliness and healthcare use using high-quality, longitudinal studies. Additionally, the association between loneliness and social care use specifically is an important future research avenue, as this was not well explored in the studies included in this review. As noted in the review by Mihalopoulos et al. [29], evaluating the economic cost of loneliness in younger populations is a clear research gap and should be considered a priority.

Future research on this topic should also include input from people with lived experience, as this review highlights that this is lacking from current studies. Future economic evaluations on this topic should consider such input to be necessary, particularly to highlight what outcomes matter most to people with lived experience. Additionally, it is key to incorporate outcome measures that are useful both in health economic evaluation (e.g. quality of life), as well as measures of loneliness and related outcomes. This is key to allow for more definitive statements to be made about the relative cost-effectiveness of different interventions, as this may be more subjective in instances where loneliness is the only common outcome measure. Finally, it is important to evaluate interventions to address wider factors like digital exclusion and poverty, to assess the potential economic impact of these in comparison to targeted loneliness interventions.

## Conclusions

Evidence relating to the cost of loneliness and the cost-effectiveness of loneliness interventions was varied and mixed. Inconsistency across findings made it difficult for any robust conclusions to be drawn about the cost of loneliness and the impacts of loneliness on health and social care use. We identified a range of different types of loneliness interventions, many of which were found to be cost-effective. However, the overall effectiveness of these interventions in reducing loneliness was often not clearly established. This review highlights a clear need for further research on this topic. Specifically, there should be an emphasis on conducting longitudinal studies with long follow-up periods and RCTs that include cost-effectiveness analyses. The cost of loneliness in younger populations and how loneliness impacts on social care across all age groups are also important research gaps that should be addressed.

## Lived experience commentary by PS, SJe and BC

Loneliness is not a unitary concept or experience and has different temporal trajectories, underlying causes, and varied pathways to potential resolution. Thus, the inconsistent findings of the review are not surprising, even with the focus on older populations. Although some reviewed papers identified participants’ sex and age, considering the impact of loneliness and interventions requires focus on discrete demographic populations, and consideration of predictable sex and age variables in likely health issues and anticipated use of services.

In reviewing the impact of loneliness on healthcare costs and usage, the bi-directional mixed findings may be explained by the issues of differential access to healthcare unique to individuals and different contexts. This might be exacerbated in some populations in future due to increasing reliance on health technology to access support. Disabilities and access challenges can contribute to social exclusion so may also lead to loneliness, although disentangling the impact of loneliness from increased need for health services may be challenging.

This systematic review addresses the subjective emotion of loneliness, but not objective measures of social isolation. As there is historic conflation of loneliness and social isolation within studies, tools and cohorts, the data may not adequately distinguish between them. The omission of linked experiences such as social isolation perhaps bypasses consideration of key underlying mechanisms for associated healthcare utilisation and costs. For example, family members may encourage seeking early support with a health concern. The growing number of older people without children, who have no family support at all, is a concern.

Few studies declared lived experience input, so we were unable to discern any accumulated wisdom from this experiential perspective regarding the health economics approaches taken. There is a need for more lived experience involvement in such studies including training offered in the technical aspects.

Loneliness, in its many forms, is unpleasant and isolating. We need ways of ameliorating it (at individual, group and societal levels), whether or not there is a quantifiable economic cost.

## Declaration of interest

None.

## Funding

This report describes independent research funded by the National Institute of Health and Care Research Policy Research Programme, conducted by the NIHR Policy Research Unit in Mental Health. The views expressed are those of the authors and not necessarily of the NIHR or the Department of Health and Social Care Research. ARG was supported by the Ramon y Cajal programme (RYC2022-038556-I), funded by the Spanish Ministry of Science, Innovation and Universities.

## Author contribution

SJo and BLE developed the original study proposal. SE drafted the protocol, with input from PB. This was reviewed and approved by PM, BLE, DM, AP, and SJo. SE led the search. SE, TS, PM, DM, PS, SJe, and ARG conducted screening. SE, HB, TS, RJ, PS, DM, PM, and ARG conducted data extraction and quality appraisal. Methodological and subject expertise was offered by PM, DM, BLE, PB, AP, MAM, and SJo. SE, PM, DM, PS, SJe, HB, TS, RJ, ARG, BC, AP, PB, BLE, and SJo contributed to the analysis and interpretation of findings. SE drafted the manuscript, which all authors reviewed, revised, and approved.

## Data availability

Data availability is not applicable to this article as no new data were created or analysed in this study.

## Supplementary materials

Supplementary file A: Search strings for each database.

Supplementary file B: Papers excluded at fulltext screening and exclusion reasons. Supplementary file C: Full quality appraisal ratings for each study.

## Notes

### Competing Interest Statement

The authors have declared no competing interest.

