## Supplementary material A for "The impact of loneliness on healthcare costs and service utilisation and the cost-effectiveness of loneliness interventions: a systematic review"

Supplementary material 1: Search strings

| Medline search (Ovid) 09/03/2023 | |
| --- | --- |
| **#** | **Search terms** |
| 1. | Loneliness.sh. |
| 2. | lonel*.ab. OR lonel*.ti. |
| 3. | "Cost-Benefit Analysis".sh. |
| 4. | "Cost-effectiveness analys?s".ab. OR "Cost-effectiveness analys?s".ti. |
| 5. | "Economic evaluation*".ab. OR "Economic evaluation*".ti. |
| 6. | "Costs and Cost Analysis".sh. |
| 7. | "Cost of Illness".sh. |
| 8. | Cost*.ab. OR Cost*.ti. |
| 9. | "Economic burden*".ab. OR "Economic burden*".ti. |
| 10. | "Economic analys?s".ab. OR "Economic analys?s".ti. |
| 11. | ("return on investment*" OR "investment return*").ab. OR ("return on investment*" OR "investment return*").ti. |
| 12. | ("service* use*" OR "service* utili?ation").ab. OR ("service* use*" OR "service* utili?ation").ti. |
| 13. | "Health Care Costs".sh. |
| 14. | #1 OR #2 |
| 15. | #3 OR #4 OR #5 OR #6 OR #7 OR #8 OR #9 OR #10 OR #11 OR #12 OR #13 |
| 16. | #14 AND #15 |
| 17. | limit #16 to yr="2008 -Current" |

| Embase search (Ovid) 09/03/2023 | |
| --- | --- |
| **#** | **Search terms** |
| 1. | Loneliness.sh. |
| 2. | lonel*.ab. OR lonel*.ti. |
| 3. | "Cost-Benefit Analysis".sh. |
| 4. | "Cost-effectiveness analysis".sh. |
| 5. | "Economic evaluation".sh. |
| 6. | "Cost of Illness".sh. |
| 7. | Cost*.ab. OR Cost*.ti. |
| 8. | "Economic burden*".ab. OR "Economic burden*".ti. |
| 9. | "Economic analys?s".ab. OR "Economic analys?s".ti. |
| 10. | ("return on investment*" OR "investment return*").ab. OR ("return on investment*" OR "investment return*").ti. |
| 11. | ("service* use*" OR "service* utili?ation").ab. OR ("service* use*" OR "service* utili?ation").ti. |
| 12. | "health care cost".sh. |
| 13. | #1 OR #2 |
| 14. | #3 OR #4 OR #5 OR #6 OR #7 OR #8 OR #9 OR #10 OR #11 OR #12 |
| 15. | #13 AND #14 |
| 16. | limit #15 to yr="2008 -Current" |

| PsycINFO search (Ovid) 09/03/2023 | |
| --- | --- |
| **#** | **Search terms** |
| 1. | Loneliness.sh. |
| 2. | lonel*.ab. OR lonel*.ti. |
| 3. | "Cost benefit analys?s".ab. OR "cost benefit analys?s".ti. |
| 4. | "Cost effectiveness analys?s".ab. OR "Cost effectiveness analys?s".ti. |
| 5. | "Economic evaluation*".ab. OR "Economic evaluation*".ti. |
| 6. | "Costs and Cost Analysis".sh. |
| 7. | Cost*.ab. OR Cost*.ti. |
| 8. | "Economic burden*".ab. OR "Economic burden*".ti. |
| 9. | "Economic analys?s".ab. OR "Economic analys?s".ti. |
| 10. | ("return on investment*" OR "investment return*").ab. OR ("return on investment*" OR "investment return*").ti. |
| 11. | ("service* use*" OR "service* utili?ation").ab. OR ("service* use*" OR "service* utili?ation").ti. |
| 12. | "Health Care Costs".sh. |
| 13. | #1 OR #2 |
| 14. | #3 OR #4 OR #5 OR #6 OR #7 OR #8 OR #9 OR #10 OR #11 OR #12 |
| 15. | #13 AND #14 |
| 16. | limit #15 to yr="2008 -Current" |

| CINAHL search (EBSCOhost) 09/03/2023 | |
| --- | --- |
| **#** | **Search terms** |
| 1. | (MH "Loneliness") |
| 2. | TI lonel* OR AB lonel* |
| 3. | (MH "Cost Benefit Analysis") |
| 4. | TI "Cost effectiveness analys?s" OR AB "Cost effectiveness analys?s" |
| 5. | TI "Economic evaluation*" OR AB "Economic evaluation*" |
| 6. | (MH "Costs and Cost Analysis") |
| 7. | (MH "Economic Aspects of Illness") |
| 8. | TI Cost* OR AB Cost* |
| 9. | TI "Economic burden*" OR AB "Economic burden*" |
| 10. | TI "Economic analys?s" OR AB "Economic analys?s" |
| 11. | TI "return on investment*" OR TI "investment return*" OR AB "return on investment*" OR AB "investment return*" |
| 12. | TI “service* use*” OR TI “service* utili?ation” OR AB “service* use*” OR AB “service* utili?ation” |
| 13. | (MH "Health Care Costs") |
| 14. | #1 OR #2 |
| 15. | #3 OR #4 OR #5 OR #6 OR #7 OR #8 OR #9 OR #10 OR #11 OR #12 OR #13 |
| 16. | #14 AND #15 |
| 17. | #14 AND #15 (Limiters - Publication Year: 2008-2023) |

| EconLit search (EBSCOhost) 09/03/2023 | |
| --- | --- |
| **#** | **Search terms** |
| 1. | lonel* |

| Google Scholar advanced search 15/05/2023 | |
| --- | --- |
| **#** | **Search terms** |
| 1. | allintitle: loneliness OR lonely; service OR cost OR economic OR healthcare OR investment (Limited: 2008-2023) |

| EThOS search 15/05/2023 | |
| --- | --- |
| **#** | **Search terms** |
| 1. | Loneliness AND cost |
| 2. | Loneliness AND service use |
| 3. | Loneliness AND investment |
| 4. | Loneliness AND economic |
| 5. | Loneliness AND healthcare |
