## Supplementary material C for "The impact of loneliness on healthcare costs and service utilisation and the cost-effectiveness of loneliness interventions: a systematic review"

Supplementary material C: full quality appraisal ratings for each study

| Quality appraisal for studies estimating the cost of loneliness using Schnitzler (2023) checklist. | | | | | |
| --- | --- | --- | --- | --- | --- |
| **Schnitzler checklist items** | ***Barnes et al., 2022*** | ***Fulton & Jupp, 2015*** | ***Meisters et al., 2021*** | ***Musich et al., 2022*** | ***Shaw et al., 2017*** |
| *Q1* | Yes | Yes | Yes | Yes | Yes |
| *Q2* | Yes | No | Yes | Yes | Yes |
| *Q3(a)* | Yes | Yes | Yes | Yes | Yes |
| *Q3(b)* | Yes | Yes | Yes | Yes | Yes |
| *Q4* | Yes | Yes | Yes | Yes | Yes |
| *Q5* | N/A | Yes | No | Yes | N/A |
| *Q6* | Yes | Yes | Yes | Yes | Yes |
| *Q7(a)* | No | No | Yes | No | Yes |
| *Q7(b)* | No | No | N/A | No | N/A |
| *Q8(a)* | Yes | Yes | Yes | Yes | Yes |
| *Q8(b)* | N/A | N/A | N/A | N/A | N/A |
| *Q9(a)* | No | Yes | No | No | Yes |
| *Q9(b)* | No | N/A | No | No | N/A |
| *Q10(a)* | Yes | Yes | Yes | Yes | Yes |
| *Q10(b)* | No | No | No | No | No |
| *Q11(a)* | No | No | No | No | Yes |
| *Q11(b)* | No | N/A | N/A | N/A | Yes |
| *Q12(a)* | No | No | No | No | Yes |
| *Q12(b)* | No | N/A | N/A | N/A | No |
| *Q12(c)* | Yes | No | Yes | No | Yes |
| *Q13* | No | Yes | Yes | No | Yes |
| *Q14* | No | No | Yes | No | No |
| *Q15* | Yes | Yes | Yes | Yes | Yes |
| *Q16(a)* | No | No | No | No | No |
| *Q16(b)* | No | No | Yes | Yes | No |
| *Q17* | No | No | No | Yes | Yes |
| ***Total*** | **10/24**  **(41.7%)** | **11/22**  **(50%)** | **14/22**  **(63.6%)** | **12/23**  **(52.2%)** | **17/22**  **(77%)** |

| Quality appraisal for healthcare use studies using Schnitzler (2023) checklist. | | | | | | | | | | |
| --- | --- | --- | --- | --- | --- | --- | --- | --- | --- | --- |
| **Schnitzler checklist items** | ***Badcock et al., 2020*** | ***Bessaha et al., 2023*** | ***Bock et al., 2018*** | ***Burns et al., 2022*** | ***Burns et al., 2021*** | ***Chamberlain et al., 2022a*** | ***Chamberlain et al., 2022b*** | ***Chen et al., 2020*** | ***Christiansen et al., 2023*** | ***Dahlberg et al., 2018*** |
| *Q1* | Yes | Yes | Yes | Yes | Yes | Yes | Yes | Yes | Yes | Yes |
| *Q2* | Yes | Yes | Yes | Yes | Yes | Yes | Yes | Yes | Yes | Yes |
| *Q3(a)* | Yes | Yes | Yes | Yes | Yes | Yes | Yes | Yes | Yes | Yes |
| *Q3(b)* | Yes | Yes | Yes | Yes | Yes | Yes | Yes | Yes | Yes | Yes |
| *Q4* | Yes | Yes | Yes | Yes | Yes | Yes | Yes | Yes | Yes | Yes |
| *Q5* | N/A | N/A | N/A | N/A | N/A | N/A | N/A | N/A | N/A | N/A |
| *Q6* | Yes | Yes | Yes | Yes | Yes | Yes | Yes | Yes | Yes | Yes |
| *Q7(a)* | Yes | No | No | No | No | No | No | No | Yes | No |
| *Q7(b)* | N/A | Yes | Yes | No | No | N/A | N/A | N/A | No | No |
| *Q8(a)* | Yes | Yes | Yes | Yes | Yes | Yes | Yes | Yes | Yes | Yes |
| *Q8(b)* | N/A | N/A | N/A | N/A | N/A | N/A | N/A | N/A | N/A | No |
| *Q9(a)* | N/A | N/A | N/A | N/A | N/A | No | No | No | No | N/A |
| *Q9(b)* | N/A | N/A | N/A | N/A | N/A | No | No | No | No | N/A |
| *Q10(a)* | Yes | Yes | Yes | Yes | Yes | Yes | Yes | Yes | Yes | No |
| *Q10(b)* | No | No | No | No | No | No | No | No | No | No |
| *Q11(a)* | N/A | N/A | N/A | N/A | N/A | N/A | N/A | N/A | N/A | N/A |
| *Q11(b)* | N/A | N/A | N/A | N/A | N/A | N/A | N/A | N/A | N/A | N/A |
| *Q12(a)* | No | No | No | No | No | No | No | Yes | Yes | No |
| *Q12(b)* | N/A | N/A | N/A | N/A | N/A | N/A | N/A | Yes | Yes | N/A |
| *Q12(c)* | Yes | No | No | Yes | Yes | N/A | N/A | Yes | No | Yes |
| *Q13* | Yes | No | No | Yes | Yes | No | No | No | Yes | No |
| *Q14* | No | Yes | Yes | Yes | Yes | Yes | Yes | Yes | Yes | Yes |
| *Q15* | Yes | Yes | Yes | Yes | Yes | Yes | Yes | Yes | Yes | Yes |
| *Q16(a)* | No | No | No | No | No | No | No | No | No | Yes |
| *Q16(b)* | No | Yes | Yes | Yes | No | No | No | No | No | No |
| *Q17* | No | No | No | No | No | No | No | No | No | Yes |
| ***Total*** | **11/21**  **(52.4%)** | **12/19**  **(63.2%)** | **12/19**  **(63.2%)** | **13/19**  **(68.4%)** | **12/19**  **(63.2%)** | **10/19**  **(52.6%)** | **10/19**  **(52.6%)** | **13/21**  **(61.9%)** | **14/22**  **(63.6%)** | **12/20**  **(60%)** |

| Quality appraisal for healthcare use studies using Schnitzler (2023) checklist (cont.). | | | | | | | | | | |
| --- | --- | --- | --- | --- | --- | --- | --- | --- | --- | --- |
| **Schnitzler checklist items** | *Denkinger et al., 2012* | *Gerst-Emerson& Jayawardhana, 2015* | *Hanratty et al., 2018* | *Lim & Chan, 2017* | *Molloy et al., 2010* | *Newall et al., 2015* | *Richard et al., 2017* | *Smith & Reed-Fitzke, 2023* | *Wang et al., 2019* | *Zhang et al., 2018* |
| *Q1* | Yes | Yes | Yes | Yes | Yes | Yes | Yes | Yes | Yes | Yes |
| *Q2* | Yes | Yes | Yes | Yes | Yes | Yes | Yes | Yes | Yes | Yes |
| *Q3(a)* | Yes | Yes | Yes | Yes | Yes | Yes | Yes | Yes | Yes | Yes |
| *Q3(b)* | Yes | Yes | Yes | Yes | Yes | Yes | Yes | Yes | Yes | Yes |
| *Q4* | Yes | Yes | Yes | Yes | Yes | Yes | Yes | Yes | Yes | Yes |
| *Q5* | N/A | N/A | N/A | N/A | N/A | N/A | Yes | N/A | N/A | N/A |
| *Q6* | Yes | Yes | Yes | Yes | Yes | Yes | Yes | Yes | Yes | Yes |
| *Q7(a)* | No | No | No | Yes | No | Yes | No | Yes | Yes | Yes |
| *Q7(b)* | N/A | No | No | N/A | No | N/A | No | N/A | N/A | N/A |
| *Q8(a)* | Yes | Yes | Yes | Yes | Yes | Yes | Yes | Yes | Yes | Yes |
| *Q8(b)* | N/A | N/A | No | N/A | N/A | N/A | N/A | N/A | N/A | N/A |
| *Q9(a)* | N/A | N/A | N/A | No | N/A | N/A | N/A | N/A | N/A | N/A |
| *Q9(b)* | N/A | N/A | N/A | No | N/A | N/A | N/A | N/A | N/A | N/A |
| *Q10(a)* | Yes | Yes | No | Yes | Yes | Yes | Yes | No | Yes | Yes |
| *Q10(b)* | No | No | N/A | No | No | No | No | N/A | No | No |
| *Q11(a)* | N/A | N/A | N/A | NA | N/A | N/A | N/A | N/A | N/A | N/A |
| *Q11(b)* | N/A | N/A | N/A | N/A | N/A | N/A | N/A | N/A | N/A | N/A |
| *Q12(a)* | No | Yes | N/A | Yes | No | No | No | No | Yes | No |
| *Q12(b)* | N/A | Yes | N/A | No | N/A | N/A | N/A | N/A | No | N/A |
| *Q12(c)* | No | Yes | N/A | Yes | No | No | Yes | Yes | Yes | Yes |
| *Q13* | No | Yes | N/A | No | Yes | Yes | No | Yes | Yes | Yes |
| *Q14* | Yes | Yes | No | Yes | No | Yes | No | Yes | Yes | Yes |
| *Q15* | Yes | Yes | No | Yes | Yes | Yes | Yes | Yes | Yes | Yes |
| *Q16(a)* | Yes | Yes | No | No | No | No | No | No | No | No |
| *Q16(b)* | No | No | No | No | No | Yes | Yes | Yes | No | No |
| *Q17* | Yes | No | Yes | No | No | No | No | No | No | No |
| ***Total*** | **12/18**  **(66.7%)** | **15/20**  **(75%)** | **8/16**  **(50%)** | **14/21**  **(66.7%)** | **10/19**  **(52.6%)** | **13/18**  **(72.2%)** | **12/20**  **(60%)** | **13/17**  **(76.5%)** | **14/19**  **(73.7%)** | **13/18**  **(72.2%)** |

| Quality appraisal for loneliness intervention studies using Evers (2005) checklist. | | | | | | | | | |
| --- | --- | --- | --- | --- | --- | --- | --- | --- | --- |
| **Evers checklist items** | *Bauer et al., 2011* | *Engel et al., 2021* | *Engel et al., 2021* | *Foster et al., 2021* | *Jones et al., 2015* | *Jones et al., 2020* | *Mallender et al., 2015* | *Mallender et al., 2015* | *Mallender et al., 2015* |
| *Q1* | Yes | Yes | Yes | Yes | Yes | Yes | No | No | No |
| *Q2* | Yes | Yes | Yes | N/A | Yes | N/A | Yes | Yes | Yes |
| *Q3* | Yes | Yes | Yes | Yes | Yes | No | Yes | Yes | Yes |
| *Q4* | Yes | Yes | Yes | Yes | No | Yes | Yes | Yes | Yes |
| *Q5* | Yes | Yes | Yes | Yes | No | Yes | Yes | Yes | Yes |
| *Q6* | Yes | Yes | Yes | No | No | No | Yes | Yes | Yes |
| *Q7* | Yes | Yes | Yes | No | No | No | Yes | Yes | Yes |
| *Q8* | Yes | No | No | No | No | No | Yes | Yes | Yes |
| *Q9* | Yes | Yes | Yes | Yes | No | Yes | Yes | Yes | Yes |
| *Q10* | Yes | Yes | Yes | NA | Yes | Yes | Yes | Yes | Yes |
| *Q11* | Yes | Yes | Yes | Yes | Yes | Yes | Yes | Yes | Yes |
| *Q12* | Yes | Yes | Yes | Yes | Yes | Yes | Yes | Yes | Yes |
| *Q13* | N/A | Yes | Yes | N/A | No | No | No | Yes | Yes |
| *Q14* | N/A | Yes | Yes | N/A | N/A | N/A | Yes | Yes | Yes |
| *Q15* | No | Yes | Yes | Yes | No | Yes | No | Yes | Yes |
| *Q16* | Yes | Yes | Yes | Yes | Yes | Yes | Yes | Yes | Yes |
| *Q17* | Yes | Yes | Yes | Yes | No | No | No | No | Yes |
| *Q18* | No | Yes | Yes | Yes | Yes | Yes | No | No | No |
| *Q19* | No | No | No | No | No | No | No | No | No |
| ***Total*** | **14/17**  **(82.4%)** | **17/19**  **(89.5%)** | **17/19**  **(89.5%)** | **12/16**  **(75%)** | **8/18**  **(44.4%)** | **10/17**  **(58.8%)** | **13/19**  **(68.4%)** | **15/19**  **(78.9%)** | **16/19**  **(84.2%)** |

| Quality appraisal for loneliness intervention studies using Evers (2005) checklist (cont.). | | | | | | | | | |
| --- | --- | --- | --- | --- | --- | --- | --- | --- | --- |
| **Evers checklist items** | *McDaid et al., 2017* | *McDaid et al., 2021* | *McDaid et al., 2021* | *McDaid & Park., 2021* | *Mountain et al., 2017* | *Simpson et al., 2014* | *Social Value Lab; 2011* | *Weiss et al., 2020* | *Willis et al., 2018* |
| *Q1* | No | Yes | Yes | Yes | Yes | Yes | No | Yes | Yes |
| *Q2* | Yes | Yes | Yes | Yes | Yes | Yes | No | Yes | N/A |
| *Q3* | Yes | Yes | Yes | Yes | Yes | Yes | Yes | Yes | Yes |
| *Q4* | Yes | Yes | Yes | Yes | Yes | Yes | Yes | Yes | Yes |
| *Q5* | Yes | Yes | Yes | Yes | Yes | No | Yes | Yes | Yes |
| *Q6* | Yes | Yes | Yes | Yes | Yes | Yes | Yes | Yes | Yes |
| *Q7* | Yes | Yes | Yes | Yes | Yes | Yes | No | Yes | Yes |
| *Q8* | Yes | Yes | Yes | Yes | Yes | Yes | Yes | No | Yes |
| *Q9* | Yes | Yes | Yes | Yes | Yes | Yes | Yes | Yes | Yes |
| *Q10* | Yes | Yes | Yes | Yes | Yes | Yes | Yes | Yes | Yes |
| *Q11* | Yes | Yes | Yes | Yes | Yes | Yes | Yes | Yes | Yes |
| *Q12* | Yes | Yes | Yes | Yes | Yes | Yes | Yes | Yes | Yes |
| *Q13* | Yes | N/A | N/A | Yes | Yes | Yes | No | Yes | N/A |
| *Q14* | Yes | No | No | Yes | No | N/A | Yes | N/A | N/A |
| *Q15* | No | No | Yes | Yes | Yes | Yes | Yes | No | No |
| *Q16* | N/A | Yes | Yes | Yes | Yes | Yes | Yes | Yes | Yes |
| *Q17* | No | Yes | No | No | Yes | Yes | Yes | No | No |
| *Q18* | No | No | No | Yes | Yes | Yes | No | Yes | Yes |
| *Q19* | No | Yes | No | No | No | Yes | No | No | No |
| ***Total*** | **13/18**  **(72.2%)** | **15/18**  **(83.3%)** | **13/18**  **(72.2%)** | **17/19**  **(89.5%)** | **17/19**  **(89.5%)** | **17/18**  **(94.4%)** | **13/19**  **(68.4%)** | **14/18**  **(77.8%)** | **13/16**  **(81.3%)** |
